# ATRIP deficiency impairs the replication stress response and manifests as microcephalic primordial dwarfism and immunodeficiency

**DOI:** 10.1101/2024.07.22.24310550

**Authors:** Evi Duthoo, Elien Beyls, Lynn Backers, Thorkell Gudjónsson, Peiquan Huang, Leander Jonckheere, Sebastian Riemann, Bram Parton, Likun Du, Veronique Debacker, Marieke De Bruyne, Levi Hoste, Ans Baeyens, Anne Vral, Eva Van Braeckel, Jens Staal, Geert Mortier, Tessa Kerre, Qiang Pan-Hammarström, Claus Storgaard Sørensen, Filomeen Haerynck, Kathleen BM Claes, Simon J Tavernier

## Abstract

ATR (Ataxia Telangiectasia and Rad3-related) kinase and its interacting protein ATRIP orchestrate the replication stress response. Two patients of independent ancestry with microcephaly, primordial dwarfism, and recurring infections were found to be homozygous for splice donor site variants of *ATRIP* exon 5, resulting in ATRIP deficiency. The c.829+5G>T patient exhibited autoimmune hemolytic anemia, lymphopenia, poor vaccine response, and intermittent neutropenia. Immunophenotyping revealed reduced CD16^+^ NK cells and absent naïve T cells, mucosal-associated invariant T cells (MAITs), and invariant natural killer T cells (iNKTs). Lymphocytic defects were characterized by T cell receptor (TCR) oligoclonality, abnormal class switch recombination (CSR), and impaired T cell proliferation. ATRIP deficiency resulted in low-grade ATR activation but impaired CHK1 phosphorylation upon genotoxic stress. Consequently, ATRIP deficient cells inadequately regulated DNA replication, leading to chromosomal instability, compromised cell cycle control, and impaired cell viability. CRISPR-Select^TIME^ confirmed reduced cell fitness induced by both variants. This study establishes ATRIP deficiency as a monogenic cause of microcephalic primordial dwarfism, highlights ATRIP’s critical role in protecting immune cells from replication stress, and brings a renewed perspective to the canonical functions of ATRIP.

## INTRODUCTION

Maintaining genomic integrity requires cells to deploy defense mechanisms to counteract various DNA-damaging assaults. The phosphatidylinositol 3-kinase-related kinases (PIKKs) ATR, ATM, and DNA-PKcs act as key mediators of the DNA damage response (DDR), initiating signaling cascades that coordinate cell cycle progression, checkpoint activation, and concurrent DNA repair^1,2^. While ATM and DNA-PKcs respond to DNA double-strand breaks (DSBs), ATR primarily safeguards DNA synthesis during the S phase^2,3^. RPA-coated single-stranded DNA (ssDNA) arises after processing of damaged DNA, including stalled replication forks, resected DSBs, and UV-induced bulky adducts, and serves as a platform for ATR activation. By phosphorylating substrates such as CHK1, ATR orchestrates the replication stress response, triggering intra-S and G2/M checkpoints, suppressing origin firing, stabilizing replication forks, and promoting replication fork restart^4–6^. ATR is essential for mammalian DNA replication and complete loss of ATR was shown to result in early embryonic lethality in mice^7–11^. A homozygous hypomorphic leaky splice variant in *ATR* (NM_001184.4 (ATR): c.2022A>G, p.(Gly674=)) was first reported by O’Driscoll et al.^12^ in two families with Seckel Syndrome^13^ (OMIM 210600), a form of microcephalic primordial dwarfism (MPD) associated with intellectual disability and distinct craniofacial features. To date, four additional ATR patients have been described, all harboring biallelic hypomorphic variants, leading to highly diminished but not abolished ATR protein levels^14–16^.

The ubiquitously expressed ATR-interacting protein (ATRIP) is an essential binding partner of ATR, illustrated by the co-dependency of ATRIP and ATR for protein stability^11^. On a functional level, ATRIP recognizes and binds RPA-ssDNA nucleoprotein filaments, allowing ATR recruitment and subsequent activation by the kinases TOPBP1 and the recently identified ETAA1^17^. The structural basis for this signaling nexus has recently been resolved: the N-terminal RPA-binding motif of ATRIP directly interacts with RPA while its coiled-coil domain is required for the simultaneous homodimerization of two ATRIP molecules. Through its C-terminal HEAT motifs, ATRIP associates with the N-terminal region of ATR and these ATR-ATRIP complexes form stable dimers of heterodimers upon recruitment to RPA-ssDNA^18–20^. Mutational analyses revealed that ATRIP-TOPBP1 interactions and downstream ATR activation depend on a region adjacent to the coiled-coil domain in ATRIP^20^. Unlike TOPBP1, which relies on both ATRIP and the RAD9–RAD1–HUS1 (9-1-1) clamp for recruitment to the ATRIP-ATR complex, ETAA1 directly interacts with RPA-ssDNA and ATR^17,21,22^. Defective ATRIP function is associated with human disease as reduced ATRIP expression was found in a patient with MPD^14^ and heterozygous *ATRIP* loss-of-function variants have been associated with breast cancer susceptibility^23^. Moreover, conditional ATRIP loss in the central nervous system in mice resulted in microcephaly, providing further evidence for a causal link between ATRIP deficiency and MPD^24^.

MPDs are a genetically heterogenous group of overlapping disorders, defined by severe microcephaly and intrauterine and postnatal growth restriction. Pathogenic variants associated with MPDs have been identified in genes involved in seemingly distinct cellular processes such as DNA replication initiation, DNA repair, cell cycle progression, and centrosome function^25–28^. Disruption of the encoded proteins consistently restrict cell proliferation dynamics, thus exposing the common biological basis of MPDs. In addition, several of the genes involved in DNA replication initiation (*MCM4, MCM10, GINS1, GINS4, POLD1-2, POLE1-2*), when disrupted, give rise to a syndromic combined immunodeficiency (CID). Although clinically variable, these immunodeficiencies have in common that they are characterized by susceptibility to severe infections by herpes viruses and defects in the natural killer (NK) cell compartment^29–37^.

Here, we report that ATRIP deficiency, caused by homozygous splice variants in *ATRIP*, manifests with features of MPD and immunodeficiency. The loss-of-function (LOF) *ATRIP* variants allow residual ATR activity, although profoundly impaired. We performed a comprehensive analysis of the functional impact of ATRIP loss on ATR signaling activity and delineated the accompanying downstream cellular outcomes. In addition to genomic instability, patient-derived cells display an inadequate ATR response to genotoxic lesions, as evidenced by dysregulated cell cycle progression, increased chromosomal sensitivity, impaired proliferation, and pronounced decline in cell viability. Modeling of the *ATRIP* variants by CRISPR-Select^TIME^ revealed reduced cell fitness^38^. Our observations highlight a more nuanced role of ATRIP within the ATR signaling pathway, clarify the biological basis of ATRIP-mediated human disease, and expand the disease phenotype associated with ATRIP deficiency.

## RESULTS

*Personally identifiable patient information was redacted in accordance with medRxiv requirements*.

### Homozygous intronic *ATRIP* variants in patients with MPD and immunodeficiency

F1Pt was born at term, small for gestational age, and presented with severe microcephaly, developmental delay, and dysmorphic features (Supplementary Fig. 1a, b). Clinical investigations revealed failure to thrive, growth retardation, grossly normal skeletal development, and a mild intellectual disability (ID). Between the ages of 0-5, she developed severe varicella and recurring respiratory tract infections (RTIs). Laboratory investigations showed lymphopenia and intermittent neutropenia (Table 1). Whole exome sequencing (WES) revealed a homozygous splice variant in intron 5 of the *ATRIP* gene (NM_13038.3): c.829+5G>T (Chr3(GRCh38):g.48457421G>T), which was confirmed by Sanger sequencing (Fig. 1a and Supplementary Fig. 1c). Importantly, WES analysis of a large cohort of patients with microcephaly identified a male patient of North Indian ancestry (patient F46.1) with another homozygous splice variant in intron 5 of *ATRIP* (c.829+2T>G)^39^. He presented with similar phenotypic features of facial dysmorphism, short stature, microcephaly, moderate ID, and recurrent RTIs (Supplementary Fig. 1a). Finally, microcephaly and short stature has already been reported in a patient with reduced ATRIP expression, due to defective splicing (patient CV1720)^14^. Reappraisal of patient CV1720’s clinical status revealed a similar phenotype of late-onset lymphopenia and intermittent neutropenia (Table 1). More detailed clinical information can be found in the case descriptions (Supplementary Materials and Supplementary Table 1).

**Table 1.** Full blood count and basic immunology screening of ATRIP patients (F1Pt and CV1720). In case of F1Pt, the timepoints represent data prior to immunoglobulin treatment (age 0-5y), pre anti-CD20 mAb treatment (age 16-20y), and post anti-CD20 mAb treatment (age 16-20y#). Blue and red bold text signify reduced and elevated values, respectively, in comparison to age-matched reference values enclosed within brackets.

| Immunological features | F1Pt |  |  | CV1720 |  |  |  |
| --- | --- | --- | --- | --- | --- | --- | --- |
|  | 0-5y | 16-20y | 16-20y# | 6-10y | 11-15y | 11-15y | 11-15y |
| **Peripheral blood count** |  |  |  |  |  |  |  |
| White blood cell counts (/μl) | 10830 (6000-15000) | **1850** (4300-9640) | **3060** (4300-9640) | 5100 (5000-15500) | **3200** (4500-13000) | **3500** (4500-13000) | 4500 (4500-13000) |
| Platelets (×10<sup>3</sup>/mL) | 311 (229-533) | **87** (175-343) | 194 (175-343) | 279 (140-400) | 218 (140-400) | 199 (140-400) | 190 (140-400) |
| Hemoglobin (g/dL) | 12.1 (11.7-15.1) | **7.4** (11.7-15.1) | 13.4 (11.7-15.1) | 13.9 (11.5-14.5) | 14.0 (13.0-18.0) | 16.0 (13.0-18.0) | 15.0 (13.0-18.0) |
| Lymphocytes (/mL) | **1191** (4000-10000) | **1308** (1230-3420) | **1050** (1230-3420) | 1870 (1500-6500) | **1430** (1500-6000) | **1190** (1500-6000) | 1830 (1500-6000) |
| Neutrophils (/mL) | 7256 (1500-7500) | **250** (1930-5780) | **1280** (1930-5780) | 2760 (1500-8000) | **1410** (1800-8000) | 1820 (1800-8000) | 2090 (1800-8000) |
| Monocytes (/mL) | 866 (400-1200) | **117** (260-780) | 410 (260-780) | 350 (200-800) | 230 (200-800) | 420 (200-800) | 330 (200-800) |
| Eosinophils (/mL) | **106** (200-600) | 161 (30-370) | 260 (30-370) | 100 (40-400) | 70 (40-400) | 70 (40-400) | 180 (40-400) |
| Basophils (/mL) | **217** (10-100) | **0** (20-80) | 40 (20-80) | 40 (20-100) | 30 (20-100) | **0** (20-100) | 20 (20-100) |
| **Lymphocyte subsets** |  |  |  |  |  |  |  |
| T cells (CD3<sup>+</sup>) (/μL) | **834** (900-4500) | 1070 (700-2100) | 1090 (700-2100) | N/A | N/A | N/A | 1060 (1000-2200) |
| Helper T cells (CD4<sup>+</sup>) (/μL) | **262** (500-2400) | 551 (300-1400) | 413 (300-1400) | N/A | N/A | N/A | 710 (530-1300) |
| Cytotoxic T cells (CD8<sup>+</sup>) (/μL) | 524 (300-1600) | 492 (200-1200) | 639 (200-1200) | N/A | N/A | N/A | **277** (330-920) |
| B cells (CD19<sup>+</sup>) (/μL) | **155** (200-2100) | 139 (100-500) | **16** (100-500) | N/A | N/A | N/A | 480 (110-570) |
| NK cells (CD56<sup>+</sup>/CD16<sup>+</sup>) (/μL) | N/A | **58.9** (90-600) | **53.5** (90-600) | N/A | N/A | N/A | 150 (70-480) |
| **Immunoglobulin levels** |  |  |  |  |  |  |  |
| IgM (g/L) | **2.2 (0.27-0.63)** | 1.51 (0.4-2.48) | **0.31** (0.4-2.48)# | N/A | N/A | N/A | N/A |
| IgA (g/L) | **1.4 (0.5-1.24)** | 1.43 (0.71-3.65) | **0.46** (0.71-3.65)# | N/A | N/A | N/A | N/A |
| Total IgG (g/L) | **21.4 (4.7-9.3)** | 15.0 (7.0-16.0) | 11.3 (7.0-16.0) | N/A | N/A | N/A | N/A |
| IgG2 (g/L) | **0.28 (0.72-3.4)** | 1.79 (1.06-6.1)* | 2.47 (1.5-6.4)* | N/A | N/A | N/A | N/A |

**Figure 1.**
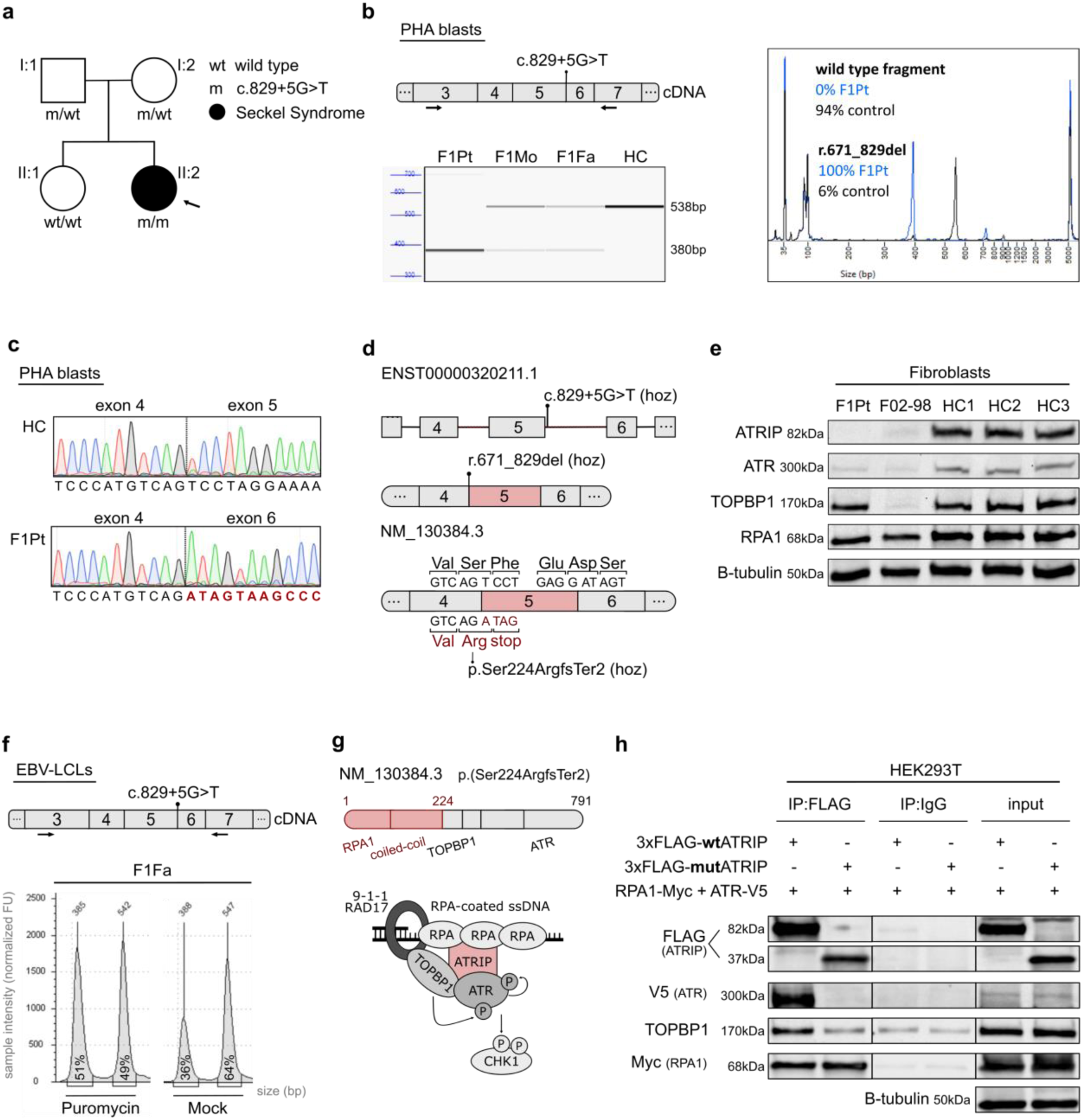
Identification of homozygous complete loss-of-function *ATRIP* variants in patients with MPD. **a** Family pedigree with allele segregation of *ATRIP* splice variant. Index patient (F1Pt) is marked with an arrow and clinical phenotype and genotype is indicated in the legend. A detailed phenotypical description can be found in Supplementary Materials. **b** Fragment analysis and size profiles of PCR-amplified cDNA extracted from PHA blasts for F1Pt, parents (F1Fa and F1Mo), and a healthy control (HC). Arrows indicate the position of forward and reverse primers used for PCR amplification. Percentages represent relative quantification of the 538bp wild type and 380bp mutant (r.671_829del) fragment. Data is representative of five independent experiments. **c** Electropherograms of cDNA extracted from PHA blasts of F1Pt and a control. Nucleotide numbering is in accordance with ENST00000320211.1. Depicted profiles are reflective of five independent experiments. **d** Schematic illustration of biallelic *ATRIP* variant effect at both transcript and amino acid level. **e** Endogenous protein expression of ATRIP and complex partners in primary fibroblasts from F1Pt, father (F1Fa), and controls (HCs). B-tubulin was used as loading control. Western blot image is reflective of four independent experiments. **f** Peak size profile in base pairs (bp) of PCR-amplified cDNA extracted from EBV immortalized lymphoblastoid cell lines (EBV-LCLs) generated from the father (F1Fa) treated with and without puromycin. Percentages represent relative ratio of wild type (835bp) and mutant (380bp) fragments. Data is reflective of two independent experiments. **g** Schematic overview of the effect of the biallelic *ATRIP* variant at the protein level and an overview of the ATR-ATRIP complex with downstream effector CHK1. **h** HEK293T cells were transiently co-transfected with 3xFLAG-tagged ATRIP (3xFLAG-wtATRIP or 3xFLAG-mutATRIP), RPA70-Myc, and ATR-V5. After immunoprecipitation with anti-FLAG or control IgG, interaction between ATRIP (FLAG) and ATR (V5), RPA70 (Myc), and endogenous TOPBP1 was examined by western blot analysis. Β-tubulin serves as a loading control. Results are reflective of three independent experiments. Source data are provided as a Source Data file.

*In silico* tools predicted inactivation of the wild-type splice donor site near exon 5 for both variants (Supplementary Table 2). Analysis of the population database gnomAD v4.0.0 revealed absence of the c.829+2T>G variant and presence of only five alleles of the c.829+5G>T variant, exclusively in a heterozygous state in the European (non-Finnish) subpopulation (MAF: 0.0004369%). Eleven homozygous variants in *ATRIP* splice boundaries, identified in gnomAD v4.0.0 (Supplementary Table 3), were present with high allele frequencies (0.0001368% - 56.0807635%) and showed exclusively low recall cutoff SpliceAI scores (recall cutoff <0.2; 0.00 - 0.18) (Supplementary Fig. 1d). Eight of the eleven homozygous variants were classified as benign (6/8) or likely benign (2/8) in ClinVar (Supplementary Table 3). Based on the clinical and genetic evidence presented above, *ATRIP* was considered a plausible candidate gene for MPD and immunodeficiency.

### *ATRIP* c.829+5G>T results in skipping of exon 5

To investigate the effect at the transcriptional level, reverse transcription PCR (RT-PCR) and Sanger sequencing was performed in PHA blasts (Fig. 1b, c) and fibroblasts (Supplementary Fig. 1e) from F1Pt. This failed to reveal a full-length transcript, but instead, showed presence of a transcript lacking exon 5 (Δex5, r.671_829del; p.(Ser224ArgfsTer2)) (Fig. 1d). In PHA blasts from her parents (F1Mo and F1Fa), both the full-length and the shorter transcript were observed, in compliance with their zygosity (Fig. 1b and Supplementary Fig. 1f). The Δex5 transcript was also detected in healthy control PHA blasts (3%) and fibroblasts (6%), indicating the presence of a naturally occurring low-abundance isoform (Fig. 1b and Supplementary Fig. 1e), consistent with the transcripts documented in the Ensembl genome browser (ENST00000635082.1). We considered the possibility that c.829+5G>T represents a leaky splice variant, however, based on targeted RNA sequencing (Supplementary Fig. 1g), RT-PCR (Supplementary Fig. 1h), and reverse transcription quantitative PCR (RT-qPCR) (Supplementary Fig. 1i), we found no evidence supporting this. Although aberrant splicing arising from c.829+2T>G was not investigated, the consequences of c.829+2T>G are expected to coincide with c.829+5G>T as it inactivates the consensus splice donor site in intron 5, demonstrated by the *in silico* splice prediction tools (Supplementary Table 2).

### Reduced ATR protein in absence of full-length ATRIP

Alternative splicing of *ATRIP* pre-mRNAs harboring c.829+5G>T or c.829+2T>G variants results in Δex5 and a subsequent shift of the reading frame (r.671_829del). At the amino acid level, this translates into a serine to arginine substitution at position 224, immediately followed by the premature stop codon TAG (p.(Ser224ArgfsTer2)) (Fig. 1d). Using western blot, we confirmed absence of full-length ATRIP in both fibroblasts and Epstein-Barr virus immortalized lymphoblastoid cell lines (EBV-LCLs) of F1Pt (Fig. 1e and Supplementary Fig. 1j, k). In absence of full-length ATRIP, ATR expression was strongly reduced in both cell types, while parental LCLs from F1Fa showed an approximately 50% reduction in both ATR and ATRIP protein levels. In line with the presumed co-dependency of ATRIP and ATR, fibroblasts from an ATR patient (F02-98)^12^ exhibited strongly reduced ATRIP levels (Fig. 1e and Supplementary Fig. 1k). In contrast, expression levels of interaction partners RPA1 and TOPBP1 were undisturbed by the absence of ATRIP (Fig. 1e and Supplementary Fig. 1j, k).

### Mutant ATRIP is loss-of-function

The premature translation stop as a consequence of Δex5 is anticipated to trigger nonsense-mediated mRNA decay (NMD). When abolishing NMD using puromycin, equal quantities of both Δex5 (±380bp, 51%) and full-length (±538bp, 49%) amplicons were observed in cDNA from EBV-LCLs of F1Fa (Fig. 1f). However, in absence of puromycin, half of the Δex5 transcript persisted (±380bp, 36% vs ±538bp, 64%), indicating that the Δex5 transcript resulting from c.829+5G>T might translate into a truncated protein. Due to the unavailability of high-quality antibodies targeting the N-terminal region of ATRIP, we confirmed the presence of a truncated protein by overexpressing 3xFLAG-tagged wild type (wt) or mutant (mut) ATRIP, which resulted in a stable 82kDa and 37kDa protein, respectively (Fig. 1h). According to the remaining amino acid sequence (AA1-224), this 37kDa ATRIP protein contains both the coiled-coil and RPA binding motif but lacks the TOPBP1 and ATR binding domains (Fig. 1g)^40^. To assess this, HEK293T cells were co-transfected with ATR-V5, RPA1-Myc, and the 3xFLAG-wtATRIP or 3xFLAG-mutATRIP constructs. Upon pulldown with an anti-FLAG antibody, immunoblotting confirmed that the 37kDa mutant was able to interact with RPA1, in accordance with earlier mutational studies^41–43^. No ATR protein could be visualized, indicating an abolished interaction. Faint TOPBP1 was detected, but at levels comparable to those of the IgG controls, suggestive of a defective interaction with TOPBP1 (Fig. 1h).

### *ATRIP* c.829+5G>T is associated with CD4^+^ T, B, and NK lymphopenia and progressive neutropenia

Given the features of immunodeficiency in patients with ATRIP deficiency, we set out to characterize the immunophenotype of F1Pt. In line with the RTIs and a severe varicella-zoster infection, F1Pt presented with reduced numbers of B and NK cells (Fig. 2a and Table 1). Low numbers of CD4^+^ T cells and an inverted CD4^+^/CD8^+^ ratio were noted (Fig. 2a and Table 1). These features were unique to F1Pt, as both the heterozygous parents and the wild-type sister had immune profiles similar to healthy controls (HCs) (Fig. 2b). Immunoglobulin substitution therapy related to IgG2 subclass deficiency and a specific pneumococcal antibody deficiency, along with azithromycin maintenance, effectively controlled the RTIs (Fig. 2c and Table 1). Over time, F1Pt developed a progressive neutropenia in presence of antineutrophil cytoplasmic antibodies, which was responsive to emergency granulopoiesis during acute infections (Fig. 2d). Between the ages of 16-20, F1Pt presented with recurrent autoimmune hemolytic anemia (AIHA) that subsided upon treatment with corticosteroids and anti-CD20 monoclonal antibody (mAb) treatment (Fig. 2e).

**Figure 2.**
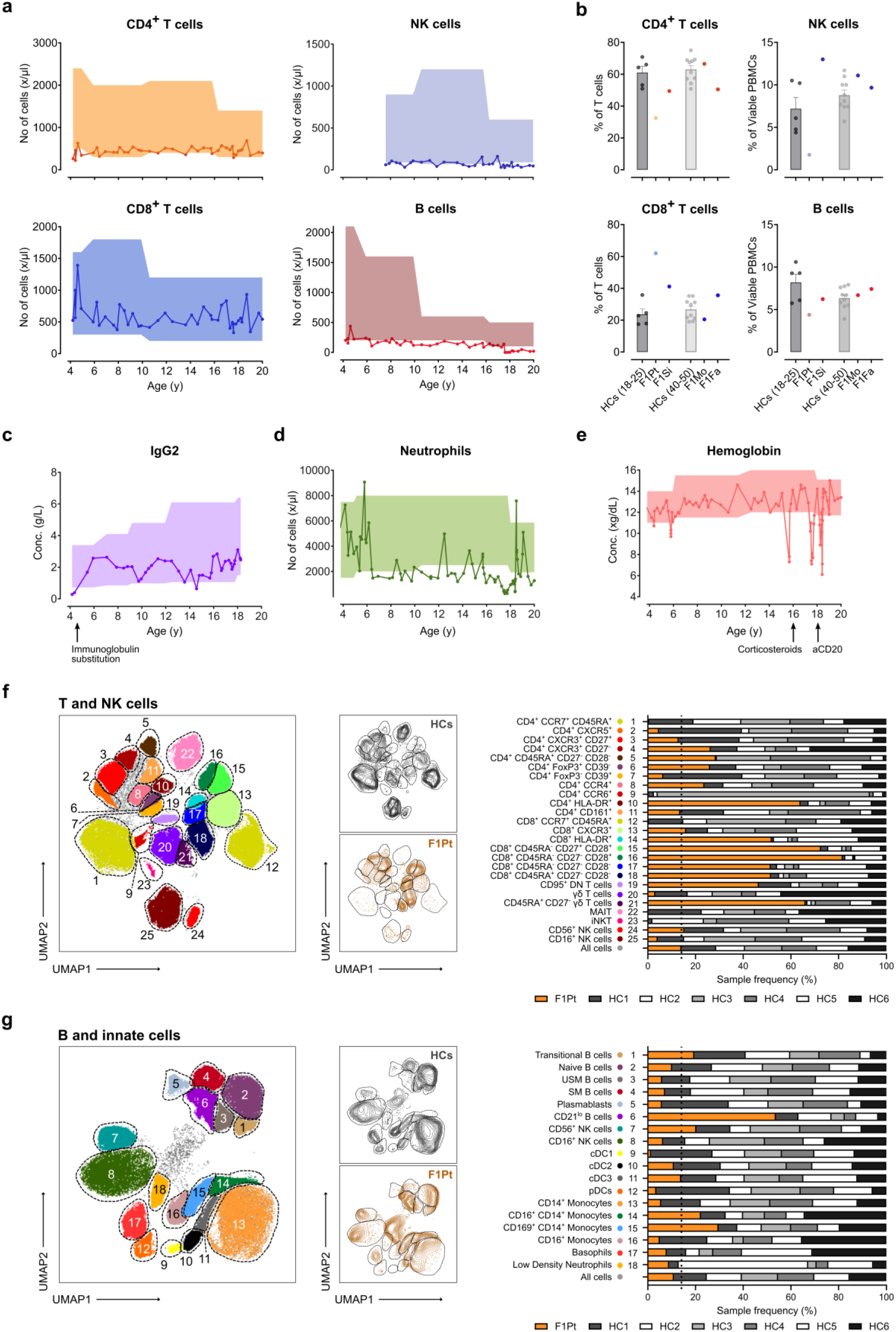
Loss of ATRIP is associated with an immune deficiency, characterized by CD4^+^ T cell lymphopenia and reduced B and CD16^+^ NK lymphocytes. **a** Total numbers (No) of CD4^+^ T, CD8^+^ T, NK, and B cells in the peripheral blood from the ATRIP patient (F1Pt) over time. Shading indicates the age-based reference range. **b** Flow cytometric (FCM) immunophenotyping of F1Pt, family members, and age-matched healthy controls (HCs). Percentages of CD4^+^ T, CD8^+^ T, NK, and B cells in PBMCs from F1Pt, sister (F1Si), mother (F1Mo), and father (F1Fa). Data represents one experiment, with each datapoint representing one biological replicate. Mean and SEM are shown. **c** IgG2 concentration (Conc.) in the peripheral blood from F1Pt over time. Shading indicates the age-based reference range. Immunoglobulin substation therapy is indicated. **d** Total numbers (No) of neutrophils in the peripheral blood from F1Pt, demonstrating intermittent neutropenia. Shading indicates the age-based reference range. **e** Hemoglobulin concentration (Conc.) in the peripheral blood from F1Pt over time. Shading indicates age-based reference range. Corticosteroid (CORT) and anti-CD20 mAb treatment (aCD20) is indicated. **f** UMAP plot depicting cluster annotation of 25 unique T and NK subsets (left). Analysis was performed using concatenated 25-parameter FCM data of PBMCs obtained from HCs (n = 6) and F1Pt. Contour plots of HCs (middle, top) and F1Pt (middle, bottom). Bar graph showing the relative proportion of HCs and F1Pt within each T and NK subset cluster (right). **g** UMAP plot demonstrating cluster annotation of 18 unique B and innate subsets (left). Analysis was performed using concatenated 25-parameter FCM data of PBMCs obtained from HCs (n = 6) and F1Pt. Contour plots of HCs (middle, top) and F1Pt (middle, bottom). Bar graph depicting the relative contribution of HCs and F1Pt within each B and innate subset cluster (right). Data is representative of one experiment (f-i). Source data are provided as a Source Data file.

### Expansion of T effector cells, low levels of CD16^+^ NK and absent MAIT, and iNKT lineages

To study the impact of ATRIP deficiency on the lymphocyte compartment in more detail, we performed high-parametric flow cytometry (FCM) (Fig. 2f, g) on PBMCs of F1Pt and age-matched HCs. Among the CD4^+^ and CD8^+^ T cells in F1Pt, we noted expansion of effector cells at the expense of naïve T cells (Fig. 2f). Especially in the CD8^+^ T cell compartment, increased percentages of HLA-DR^+^ and CD27^−^ CD28^−^ T cells were noted, indicating ongoing activation and exhaustion. Similarly, CD95^+^ double-negative (DN) T cells were expanded. In addition to diminished naïve T cell levels, we noted the absence of both mucosal-associated invariant T (MAIT) and invariant natural killer T (iNKT) cells. The observed NK lymphopenia was characterized by a selective reduction of CD16^+^ NK cells (Fig. 2f). NK cytotoxic potential was assessed and revealed normal CD107a upregulation upon stimulation with K652. In line with the higher proportion of CD56^bright^ NK cells in F1Pt, perforin expression was reduced (data not shown). In a similar approach, the B cell and innate compartment was investigated in detail (Fig. 2g). CD21^lo^ B cells were expanded in F1Pt at the expense of CD21^hi^ memory B cells and plasmablasts. In the innate compartment, we noted lower percentages of type 1 dendritic cells (DC1) and plasmacytoid (p)DCs. Among monocyte subsets, both inflammatory and CD169^+^ expressing monocyte subsets were increased. Manual gating of the FCM data confirmed the main findings of this analysis (Supplementary Fig. 2a-c).

### TCR oligoclonality and altered antibody class switch recombination in absence of ATRIP

Given the diminished levels of naïve T cells and absence of MAIT cells in F1Pt, we looked for potential disparities in the recombinational processes of antigen receptors. To study this in more detail, CITE-seq (cellular indexing of transcriptomes and epitopes by sequencing) combined with T cell receptor (TCR) and B cell receptor (BCR) sequencing of PBMCs, collected prior to anti-CD20 mAb treatment, was performed. CITE-seq profiling confirmed the observed immune defects (Fig. 3a, and Supplementary Fig. 3a). By visualizing both the usage and pairing of unique *TRAV* and *TRBV* genes, we observed a decreased diversity in the TCR repertoire in F1Pt compared to HCs (Fig. 3b, c and Supplementary Fig. 3b, c). Oligoclonality was most evident in CD8^+^ effector memory (TEM) cells, suggesting clonal expansion (Fig. 3b, c). Using a similar analysis based on *IGH* transcripts, we observed no restrictions in BCR repertoire diversity (Supplementary Fig. 3e, f). Changes in complementarity-determining region 3 (CDR3) length and composition have been described in immunodeficiencies^44,45^. Comparing the CDR3 length profiles of *TRAV*, *TRBV,* and *IGH* transcripts, we observed no consistent differences between F1Pt and HCs (Fig. 3d and Supplementary Fig. 3d, g). These findings are indicative of competent V(D)J recombination, suggesting adequate repair by non-homologous end-joining (NHEJ) in the absence of ATRIP.

**Figure 3.**
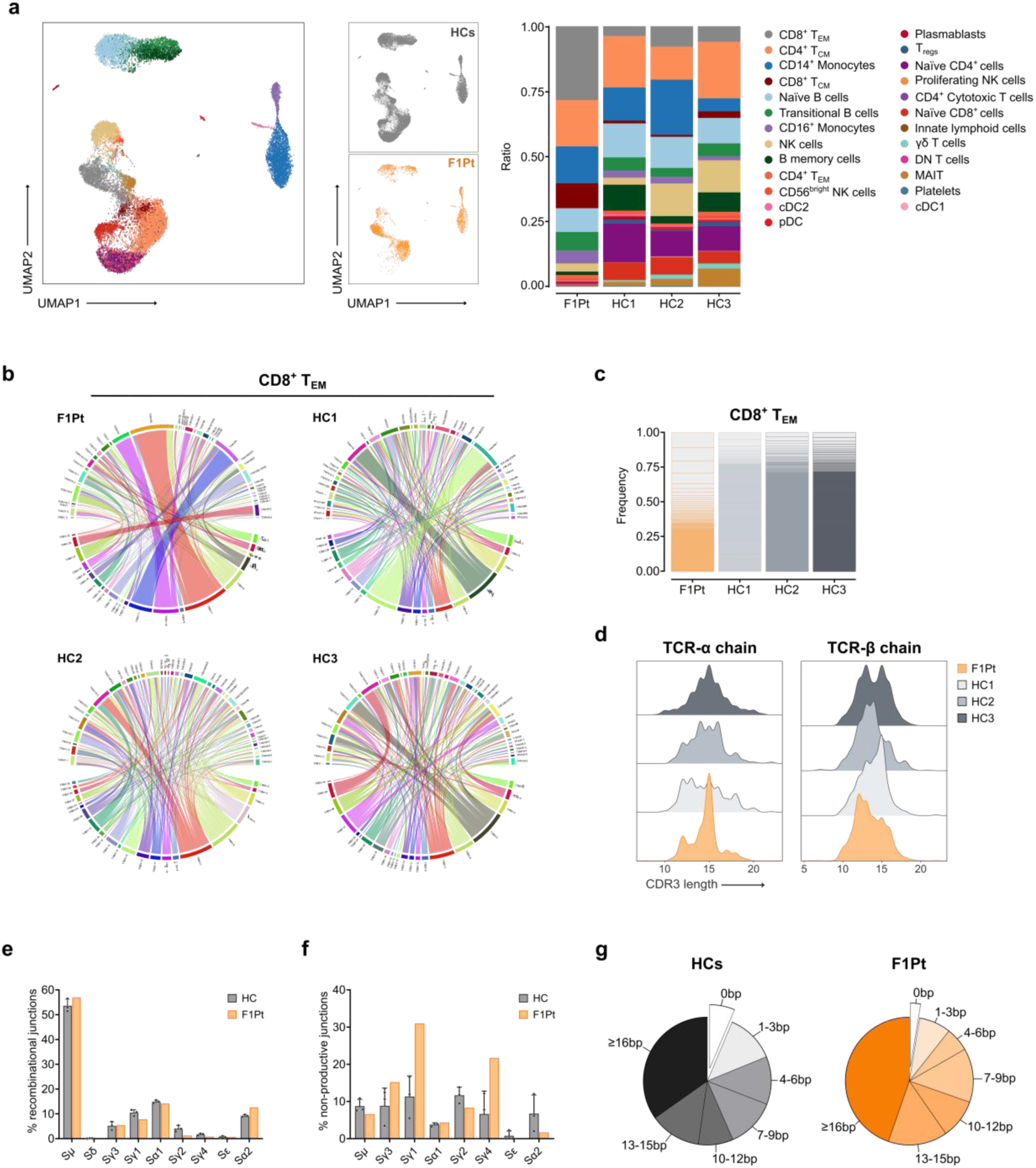
TCR oligoclonality in CD8^+^ T_EM_ cells and altered class switch recombination (CSR) in absence of ATRIP. **a** CITE-seq profiling of PBMCs from healthy controls (HCs) (n =3) and ATRIP patient (F1Pt), identifying 25 immune subsets. UMAP visualization of pooled CITE-seq data of HCs and F1Pt, displaying the identified subsets (left). UMAP plot of HCs (middle, top) and F1Pt (middle, bottom). Ratio of the 25 subsets in HCs and F1Pt, ranked based on the prevalence in F1Pt (right). **b** Circos plots showing the *TRBV* and *TRAV* pairing pattern of CD8^+^ T effector memory (T_EM_) cells of healthy controls (HCs) and ATRIP patient (F1Pt). **c** Frequency of unique CD8^+^ T_EM_ cell clones in HCs and F1Pt. **d** Distribution of the CDR3 region lengths of TCR-α and TCR-β clones from HCs and F1Pt CD8^+^ T_EM_ cells. **e** Frequency of class switch recombination junctions per S region in HCs (n = 3) (junctions; n = 8305) and F1Pt (junctions; n = 2758). Mean and SD are shown. **f** Proportion of non-productive junctions (inversional recombination) per S region in HCs (n = 3) (junctions; n = 652) and F1Pt (junctions; n = 225). Mean and SD are shown. **g** Pie charts demonstrating the microhomology usage at Sμ-Sα junctions in HCs (n = 3) and F1Pt. Data represents one experiment (a-f). Source data are provided as a Source Data file.

Affinity maturation and isotype switching of antibodies require efficient DNA recombination processes called somatic hypermutation and class switch recombination (CSR), respectively. A potential role for ATR signaling in CSR has been suggested in previous studies, owing to its regulatory function in DNA replication and proliferation^15,46,47^. Using a modified version of linear amplification-mediated high-throughput genome-wide translocation sequencing (LAM-HTGTS), we assessed CSR patterns in ATRIP deficient PBMCs^48,49^. Our results revealed an altered isotype switching profile in F1Pt, with decreased proportions of recombination between Sµ and Sγ1, Sγ2, and Sγ4 (Fig. 3e). A marked increase in inversional Sµ-Sγ1 and Sµ-Sγ4 junctions was noted, indicating reduced efficiency in IgG subclass switching (Fig. 3f). We observed increased usage of longer microhomologies (≥10 bp) at the Sµ-Sα switch junctions in F1Pt as compared to HCs (Fig. 3g), indicating increased repair via alternative end joining. Given that alternative end joining depends on DNA end resection during early S phase, these results are suggestive of disruptions in cell cycle dynamics^50^.

### *ATRIP* c.829+5G>T presents with a DNA repair signature and interferon-driven immune dysregulation

We performed Gene Set Enrichment Analysis (GSEA) to examine the differentially expressed genes in PBMCs of F1Pt compared to HCs. Among the most altered biological processes (adj P<0.05; Log2: ±1), we noted a distinct DNA repair signature in PBMCs from F1Pt (Fig. 4a, b and Supplementary Fig. 4a, b), which was also evident in both T effector and NK subsets (Supplementary Fig. 4c, d). Likely related to F1Pt’s autoimmune phenotype, an upregulated interferon (both type I and II) response was observed. Analysis of cytokine expression in serum samples collected during an episode of AIHA and after successful treatment with anti-CD20 mAb confirmed increased interferon signaling (IFNγ and IP10) and indicated both innate and T lymphocyte subset activation (IL-10, IL-18, IL-27, IL-31) (Fig. 4c). While treatment of AIHA with corticosteroids and anti-CD20 mAb resulted in partial reduction of all cytokines, the Th2-signature cytokines IL-10 and IL-31 persisted. FCM analysis confirmed the response to anti-CD20 mAb therapy, with partial normalization of interferon-regulated proteins CD64 and CD169 on classical monocytes (Fig. 4d). While effectively targeting B cells, anti-CD20 mAb treatment did not normalize NK and T cell subset percentages, nor did it normalize T cell maturation (Supplementary Fig. 4e-g). Additionally, treatment had minimal effect on T cell activation, as indicated by increased levels of ICOS, OX40, and PD1 (Supplementary Fig. 4h).

**Figure 4.**
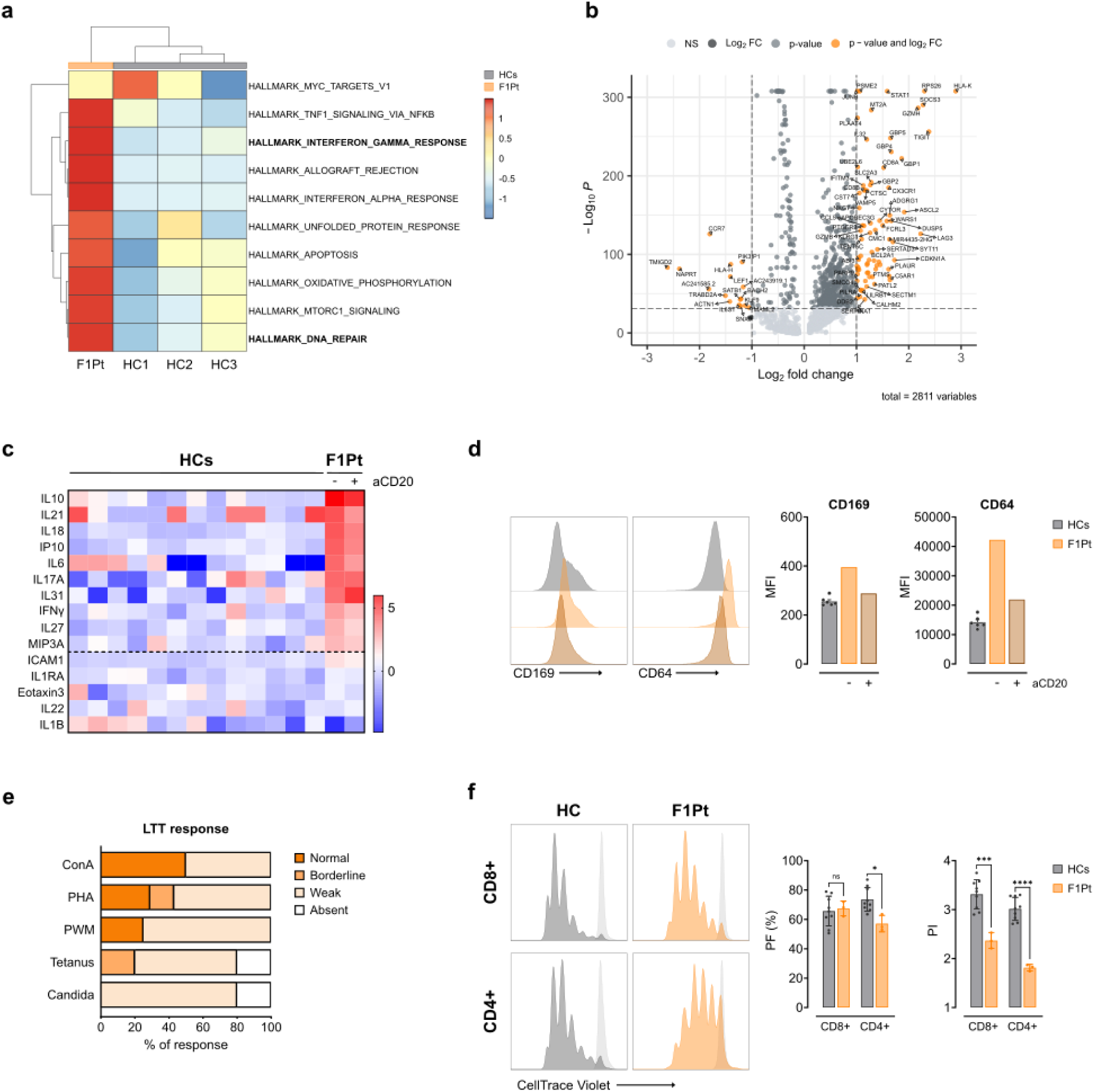
ATRIP deficiency presents with a DNA repair signature, interferon-driven immune dysregulation, and impaired T cell proliferation. **a** Heatmap representing the top 10 enriched hallmark gene sets (MSigDB) in PBMCs of F1Pt compared to HCs (n = 3). **b** Volcano plot showing the differentially expressed genes in PBMCs of F1Pt compared to HCs (n = 3). **c** Heatmap displaying the serum concentration of the cytokines IL-10, IL-21, IL-18, IP-10, IL-6, IL-17A, IL-31, IFNγ, IL-27, MIP31, ICAM-1, IL-1RA, Eotaxin-3, IL-22, and IL-1β in HCs (n = 13) and F1Pt pre- and post-treatment with anti-CD20 mAb. **d** CD169 and CD64 expression on CD14^+^ monocytes of HCs (n = 6) and F1Pt pre- and post-treatment with anti-CD20 mAb (aCD20). Histograms of a representative HC and F1Pt are shown. Bar plots depict median fluorescence (MFI). Mean and SEM are shown. **e** Lymphocyte transformation test (LTT) demonstrating the proliferative reponse of F1Pt whole blood to various stimuli. Data shows at least two independent analyses. **f** Proliferation analysis of CD8^+^ and CD4^+^ PHA blasts from HCs (n = 9) and F1Pt. PBMCs were labeled with CellTrace Violet (CTV) and stimulated with PHA for 96h. Precursor frequency (PF) and proliferation index (PI) of three independent experiments are depicted, with each datapoint representing one biological replicate. Mean and SD are shown. ns: not significant, *p<0.05, ***p<0.001, **** p<0.0001 (multiple unpaired t-tests). Source data are provided as a Source Data file.

### Impaired T cell proliferation in absence of ATRIP

Considering that GSEA and CSR analyses indicated dysregulated DNA repair and cell cycle dynamics in F1Pt cells, we proceeded to investigate the proliferative capacity of T cells in more detail. The lymphocyte transformation test (LTT) performed on whole blood of F1Pt revealed reduced or absent T cell proliferation upon stimulation with both mitogens and specific antigens (candida, tetanus), respectively (Fig. 4e). Using a more comprehensive study determining both the precursor frequency (PF) and proliferation index (PI) of CellTrace Violet (CTV) labeled T cells, we noted a substantial reduction in the proliferation capacity of both CD4^+^ and CD8^+^ T cells of F1Pt upon stimulation with the mitogen PHA (Fig. 4f). In conclusion, the immunophenotype of ATRIP deficiency is characterized by defects in all three lymphocytic lineages, an interferon-driven immune dysregulation, and a profound T cell proliferative defect.

### Loss of ATRIP does not impair ATR recruitment and auto-phosphorylation

Next, we set out to characterize the molecular processes driving disease in absence of ATRIP. As localization of ATR to DNA damage sites is a prerequisite for ATR activation, we assessed nuclear ATR foci by immunofluorescence. Unexpectedly, this revealed that F1Pt fibroblasts were competent in forming ATR foci in response to the known ATR-activating DNA damage inducers mitomycin C (MMC), a DNA interstrand crosslinker (ICL), UV, and ionizing radiation (IR). In addition, F02-98 fibroblasts (ATR patient) showed normal recruitment of ATR (Fig. 5a and Supplementary Fig. 5a), despite substantially reduced total ATR protein levels (11% of HC levels; Supplementary Fig. 1g). No significant differences in foci number per cell were observed between HC and F1Pt fibroblasts following MMC and IR treatment, whereas UV exposure resulted in a slightly higher amount of ATR foci in F1Pt and F02-98 fibroblasts. The higher number of spontaneous foci in mock conditions suggests persistent DNA damage (Fig. 5a and Supplementary Fig. 5a).

**Figure 5.**
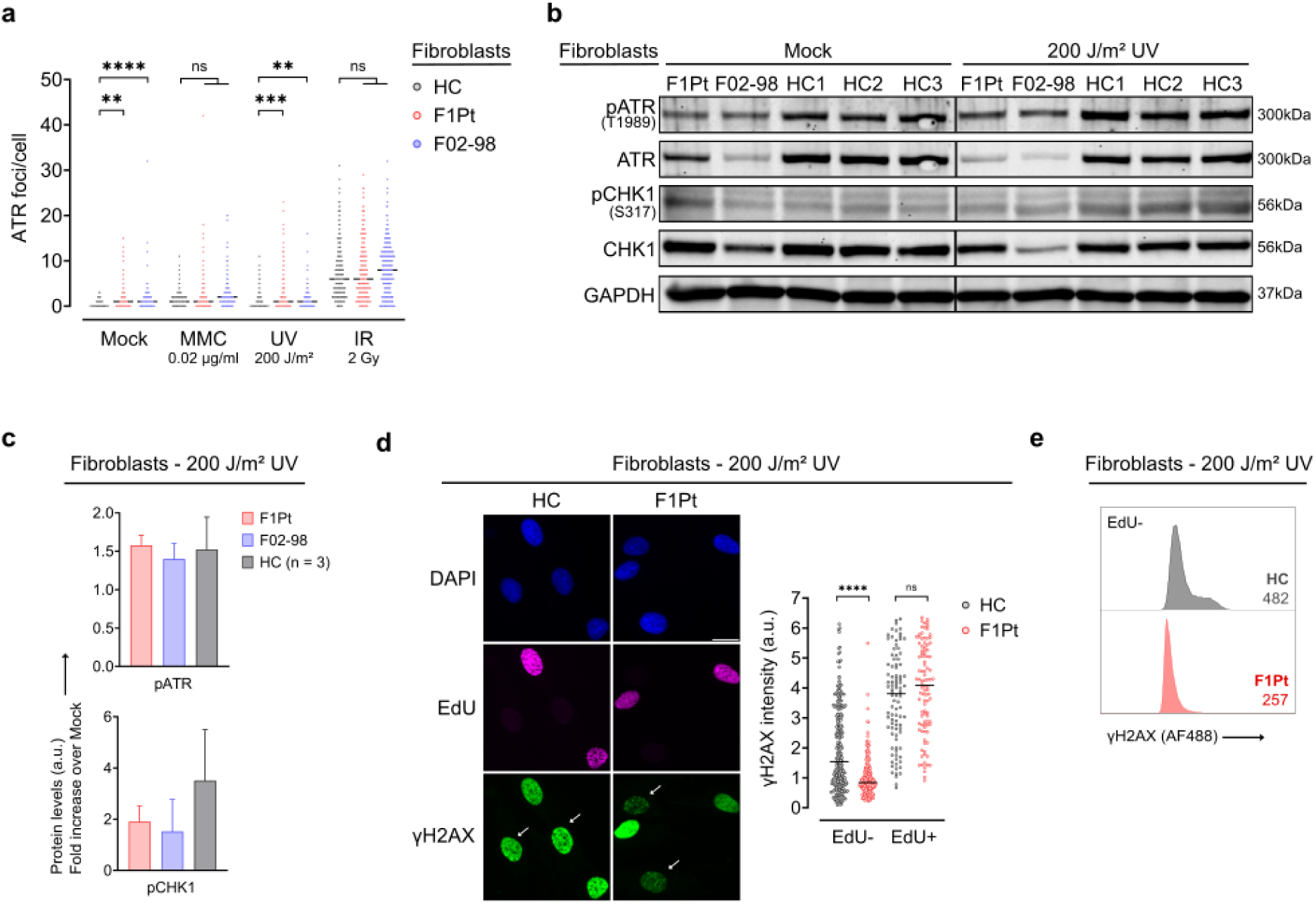
Absence of ATRIP does not abolish recruitment of ATR and its ability to phosphorylate substrates but reveals an insufficient ATR signaling response. **a** Fibroblasts from a healthy control (HC), ATRIP patient (F1Pt) and ATR patient (F02-98) were left untreated or exposed to 0.02 µg/ml Mitomycin C (MMC), 200 J/m² UV or 2 Gy IR. ATR was stained by immunofluorescence after 24h (MMC) or 3h (UV and IR) after exposure and ATR nuclear foci were quantified. Dot plot represents pooled data from three independent experiments; at least 150 cells were analyzed for each condition. The median number of foci is depicted. ns: not significant, **p<0.01, ***p<0.001, ****p<0.0001 (Kruskall-Wallis test and Dunn’s multiple comparisons test). **b** Protein expression of phosphorylation events (T1989-pATR, S317-pCHK1) and total protein (ATR, CHK1) 3h post 200 J/m² UV radiation. Immunoblotting was performed on fibroblasts from F1Pt, F02-98, and HCs (n = 3). Western blot is representative of three independent experiments. GAPDH serves as a loading control. **c** pATR and pCHK1 levels shown in 2B were quantified. Bar graph depicts pATR and pCHK1 levels post UV treatment, expressed as a fold increase over the levels observed in the Mock condition. Mean and SD are shown. **d** Quantification of yH2AX nuclear fluorescence in HC and F1Pt fibroblasts 3h after 200 J/m² UV exposure and concomitant EdU pulse-labeling. The mean yH2AX intensity per nucleus is shown for EdU- and EdU+ fibroblasts. Dot plot represents pooled data from three independent experiments. At least 200 (EdU-) or 90 (EdU+) cells were analyzed per condition. The median value is depicted. ns: not significant, ****p<0.0001 (multiple Mann-Whitney tests and Bonferroni-Dunn multiple comparisons test). Representative immunofluorescence images with DAPI, EdU, and yH2AX staining is shown (left). White arrows indicate EdU-cells. Scale bars are 20 µm. **e** yH2AX expression was determined by flow cytometric analysis 3h following 200 J/m² UV exposure in EdU-(G0/G1 and G2/M phase) fibroblasts of HC and F1Pt. Median fluorescence intensity (MFI) of yH2AX (AF488) is annotated on the histogram. Data are reflective of one experiment. Source data are provided as a Source Data file.

Next, ATR-dependent substrate phosphorylation was investigated as a measure for ATR activation. Autophosphorylation of ATR on residue T1989 has been identified as a hallmark of its active state and is regulated by DNA damage induction^51,52^. T1989 phosphorylation (hereafter referred to as pATR) was readily detected in untreated F1Pt and F02-98 cells and further increased upon UV treatment, although absolute pATR levels were reduced compared to control fibroblasts (Fig. 5b and Supplementary Fig. 5b). Interestingly, when expressing UV-induced pATR levels as a fold increase over basal levels, a similar 1.5-fold induction was observed for F1Pt, F02-98, and control cells (Fig. 5c). Of note, the specificity for T1989 phosphorylation was confirmed using lambda phosphatase (λPPase) treatment (Supplementary Fig. 5c). Full activation of CHK1, the major downstream effector of the ATR signaling pathway, requires phosphorylation by ATR ^6^. Following UV exposure, CHK1 phosphorylation on residue S317 (indicated as pCHK1) was reduced in both F1Pt and F02-98 fibroblasts compared to HCs (Fig. 5b and Supplementary Fig. 5b). This reduction was also evident when the fold induction over basal levels was quantified (Fig. 5c). Of note, total CHK1 levels were reduced in F02-98, but not in F1Pt fibroblasts (Fig. 5b), a possible consequence of the high passage number and senescence of the F02-98 cells^53^.

DNA repair intermediates formed after UV irradiation trigger H2AX phosphorylation at S139 (known as γH2AX), a process mainly mediated by ATR and found to be impaired in ATR patient cells^12,15,54,55^. Following UV exposure, a strong pan-nuclear γH2AX positivity was readily noted via immunofluorescence in control fibroblasts (Fig. 5d). In contrast, γH2AX was almost entirely absent in EdU-nuclei of F1Pt fibroblasts, while it was retained in EdU+ nuclei. Consistent with this, FCM analysis of γH2AX expression in EdU-cells showed decreased MFI values in F1Pt (Fig. 5e). In conclusion, while the exact mechanism of ATR recruitment and activation remains unclear, our observations indicate that ATRIP is dispensable for the initial recruitment and subsequent autophosphorylation of ATR. Downstream ATR substrate phosphorylation was impaired but not abolished, providing evidence for an inadequate ATR signaling response in absence of ATRIP.

### Compromised replication stress response and cell cycle progression in absence of ATRIP

Replication stress induces stalling of replication forks and exposes single-stranded DNA (ssDNA) that is rapidly coated with RPA. Subsequent ATR activation safeguards genomic stability and prevents further fork stalling and collapse by restraining fork progression and suppressing origin firing. In addition, the ATR kinase is responsible for stabilizing stalled forks, thereby facilitating fork restart upon resolved stress. We aimed to thoroughly define the *in vitro* phenotype associated with ATRIP deficiency by investigating the cellular consequences of genotoxic treatments.

Previous studies noted comparable cell cycle distributions in untreated cells of ATR patients and HCs^14,15^. Consistent with this, we observed no differences in cell cycle ratios between F1Pt and control fibroblasts or PHA blasts (Fig. 6a, b and Supplementary Fig. 6a). To investigate the replication stress response, we treated F1Pt fibroblasts with MMC and assessed the replicative response using FCM (Fig. 6a). Whereas HC cells efficiently suppressed origin firing and fork progression upon MMC treatment (indicated by reduced EdU intensity), F1Pt cells showed no substantial response (Fig. 6c). To verify ATR involvement, we pretreated cells with an ATR inhibitor (ATRi). Upon ATRi treatment, the EdU intensity of HC cells mimicked that of ATRIP deficient cells, confirming impaired ATR signaling in F1Pt cells. Additionally, DAPI histograms of S phase cells showed accumulation of HC cells in late S phase following MMC, in contrast to F1Pt cells that retained a diffuse DAPI profile similar to non-treated cells.

**Figure 6.**
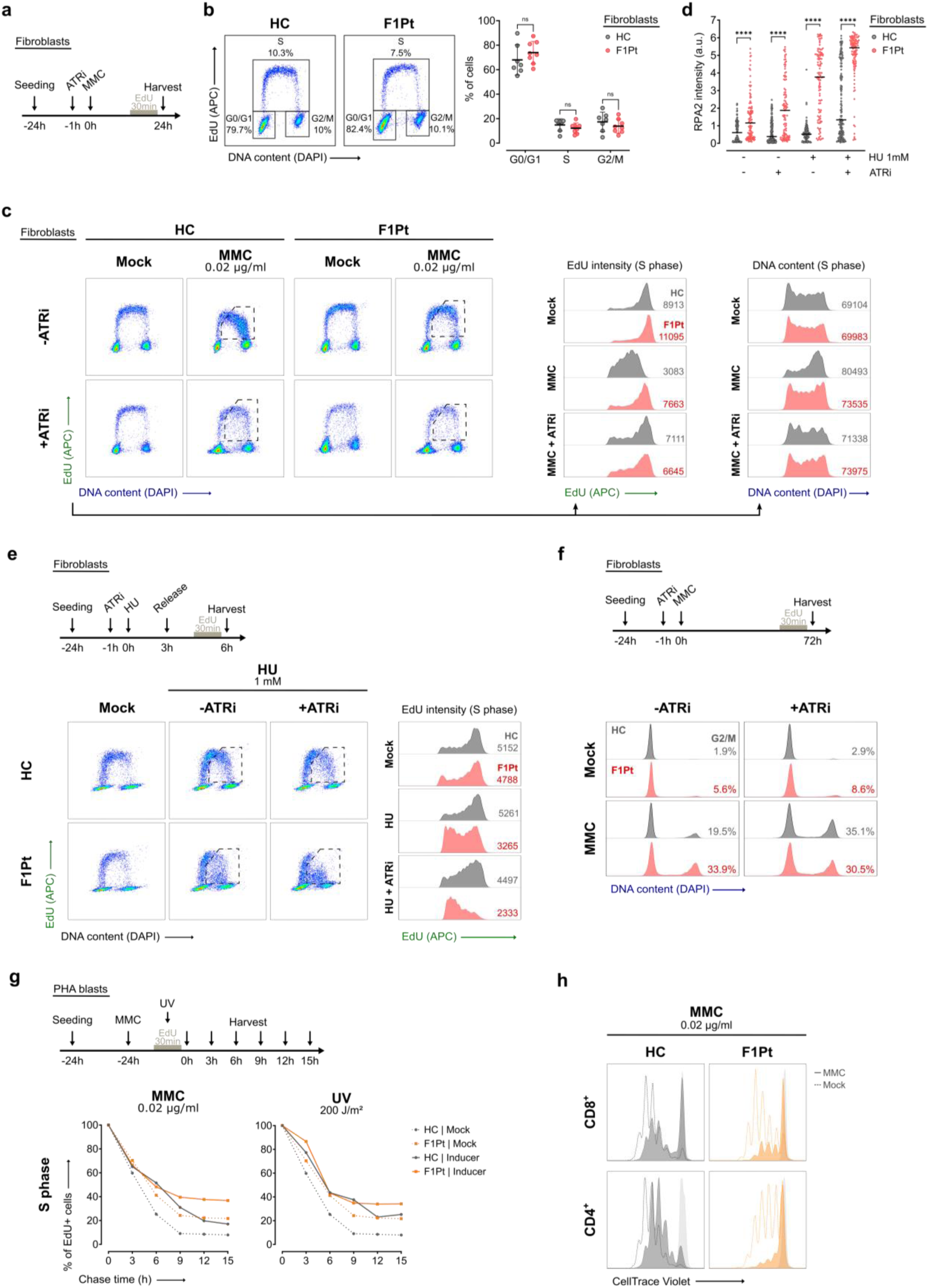
Loss of ATRIP results in compromised DNA replication and impaired cell cycle progression following replication stress. **a** Schematic outlining the treatment protocol of the flow cytometric (FCM) assay used in **b** and **c**. **b** Cell cycle distributions of untreated fibroblasts from a healthy control (HC) and ATRIP patient (F1Pt). Representative FCM plots are shown (left). Scatter dot plot depicts data from eight independent experiments (right). Mean and SD are shown. ns: not significant (multiple paired t-tests). **c** FCM EdU pulse-labeling profiles of Mitomycin C (MMC) treated HC and F1Pt fibroblasts (top). Cells were exposed to 0.02 µg/ml MMC for 24h in the absence or presence of an ATR kinase inhibitor (ATRi, 20 nM). Histograms of EdU and DAPI intensity in S phase cells are shown (bottom). The median fluorescence intensity (MFI) is annotated on the histogram. Data is representative of three independent experiments. **d** Quantification of RPA nuclear fluorescence in HC and F1Pt fibroblasts 3h after 1 mM Hydroxyurea (HU) exposure and concomitant EdU pulse-labeling. The mean RPA intensity per nucleus is shown for EdU+ cells. Dot plot represents data from pooled data from three experiments. The median value is depicted. ns: not significant; ***p<0.001 (multiple Mann-Whitney tests and Bonferroni-Dunn multiple comparisons test). **e** FCM EdU pulse-labeling profiles of HC and F1Pt fibroblasts after the release from HU treatment (left). Cells were exposed to 1 mM HU for 3h, released for 3h, and subsequently harvested. Histograms of EdU intensity in S phase cells are shown (right). Data are representative of two experiments. **f** Cell cycle profiles of HC and F1Pt fibroblasts following 72h of 0.02 µg/ml MMC treatment, with and without 20 nM ATRi. The percentage of cells in G2/M phase is indicated. Data is representative of three independent experiments. **g** EdU pulse-chase kinetics of HC and F1Pt PHA blasts. Cells were untreated or treated with genotoxic inducers (0.02 µg/ml MMC or 200 J/m² UV), pulse-labeled with EdU, and harvested at indicated time points. Kinetic plots show percentages of EdU+ cells present in S phase (middle) and is representative of three independent experiments. **h** CellTrace Violet (CTV) profiles of CD8^+^ and CD4^+^ PHA blasts from HC and F1Pt after 96h of culture in the presence or absence of 0.02 µg/ml MMC. Data is representative of two experiment. Source data are provided as a Source Data file.

In absence of timely ATR activation, excessive origin firing results in accumulation of RPA-ssDNA nucleoprotein filaments, progressively depleting the nuclear pool of RPA and preceding compromised fork integrity^56^. Using immunofluorescence, we observed significantly higher RPA2 intensity in S phase F1Pt fibroblasts compared to HCs, both in mock and upon hydroxyurea (HU) treatment (Fig. 6d and Supplementary Fig. 6b). HU induces widespread fork stalling through dNTP depletion, as evidenced by the complete loss of EdU incorporation after 3h of HU treatment (Supplementary Fig. 6c)^57^. Additional treatment with ATRi further increased RPA2 intensity in both F1Pt and HCs, although the RPA2 intensity levels in HCs did not reach those observed in F1Pt.

Next, we determined whether patient fibroblasts retained their ability to recover from acute HU-induced replication stress. While HC cells readily resumed DNA synthesis, F1Pt fibroblasts demonstrated reduced levels of fork recovery following replication fork stalling, indicated by decreased EdU incorporation following HU release. Combined HU and ATRi treatment impeded replication recovery in HC cells and further intensified the observed impairment in F1Pt cells (Fig. 6e).

Regulation of cell cycle progression is crucial to allow repair of damaged DNA and completion of DNA replication prior to mitosis. Pronounced G2/M arrest has been described as a hallmark of ATR deficiency, reflecting the direct consequence of acquired DNA damage during the S phase^56^ Accordingly, F1Pt fibroblasts and PHA blasts accumulated in the G2/M phase after MMC exposure (Fig. 6f and Supplementary Fig. 6a). ATRi treatment in HC cells resulted in similar checkpoint activation, confirming that the observed G2/M arrest is a direct result of defective ATR signaling in the S phase (Fig. 6f). An additional established function of ATR is the G2/M checkpoint activation in response to IR, preventing premature mitotic entry in the presence of severely damaged DNA (Supplementary Fig. 6d)^58,59^. In ATRIP deficient cells, the ATR signaling pathway retained sufficient activity to initiate G2/M checkpoint arrest upon irradiation, albeit at a significantly lower level compared to HC cells. (Supplementary Fig. 6d).

As our data did not indicate significant deviations in the cell cycle distribution of unperturbed patient cells, we extended our investigations to assess the S phase kinetics using an EdU pulse-chase assay (Fig. 6g and Supplementary Fig. 6e)^60^. We found that unperturbed F1Pt PHA blasts displayed a prolonged S phase, compared to HC cells. Exposure to MMC and UV increased the S phase-delay in both F1Pt and HC cells, with a more pronounced effect in F1Pt cells. In light of these findings, we speculated that proficient ATR signaling is essential for facilitating T cell proliferation under conditions of excessive replicative stress. Indeed, upon treatment with MMC, we observed a strongly diminished proliferative response and subsequent division rate of CTV-labeled T cells of F1Pt (PF and PI, respectively), particularly in CD4^+^ T cells (Fig. 6h and Supplementary Fig. 6f).

Taken together, defective ATR signaling in ATRIP deficient cells results in an impaired replication stress response, consequently compromising cell cycle progression and proliferation.

### *ATRIP* c.829+5G>T cells display DNA damage, chromosomal sensitivity, and impeded survival

The consequences of compromised replication and subsequent accumulation of DNA damage typically manifest post-mitosis, resulting in chromosomal breakage and cell death. To assess spontaneously occurring DNA damage, we examined the formation of γH2AX nuclear foci, a well-established marker of DNA double-strand breaks (DSBs). Quantification of γH2AX in EdU-cells revealed a significantly higher number of foci in F1Pt and F02-98 fibroblasts (Fig. 7a and Supplementary Fig. 7a). To evaluate DSB repair fidelity, we analyzed micronucleus (MN) formation using 3 different assays: the G0 MN assay, the MMC MN test, and a newly developed S MN assay (Fig. 7b). F1Pt PHA blasts showed increased spontaneous MN levels with the S MN assay, opposed to the normal levels observed with the G0 MN test (Fig. 7c). This discrepancy suggests that DNA damage and subsequent chromosomal breakage only becomes apparent after multiple cell cycles have been completed. Correspondingly, immunofluorescence preparations of proliferating fibroblasts showed higher MN frequencies in F1Pt and F02-98 compared to HCs (data not shown).

**Figure 7.**
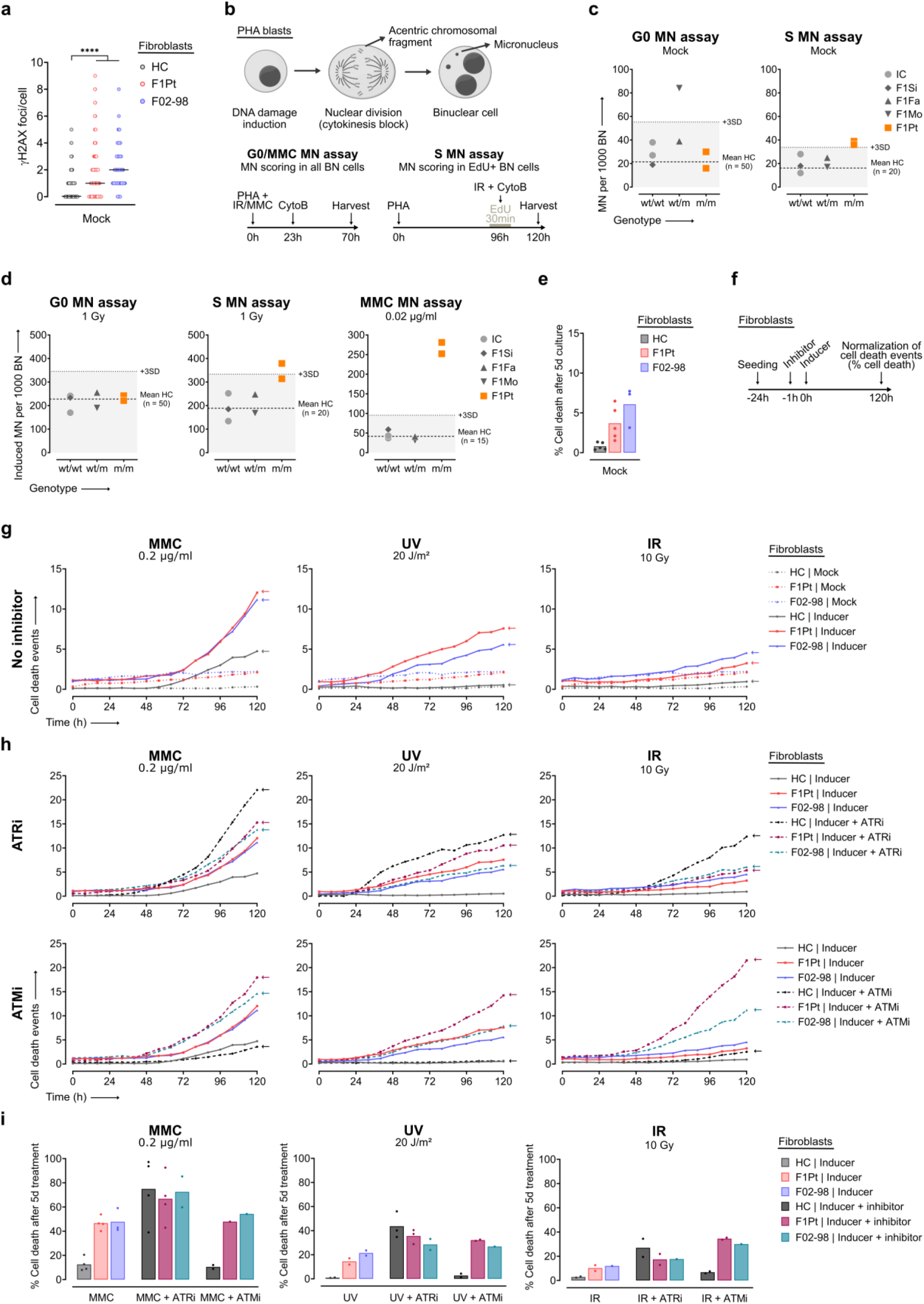
Increased spontaneous DNA damage, chromosomal sensitivity, and impaired survival upon exposure to genotoxic inducers requiring ATR pathway activation in Seckel syndrome cells. **a** Immunofluorescence analysis of yH2AX foci in untreated EdU-fibroblasts from a healthy control (HC), ATRIP patient (F1Pt), and ATR patient (F02-98). yH2AX nuclear foci were quantified and are shown in a dot plot representing pooled data from three independent experiments. At least 150 cells were analyzed for each condition. The median number of foci is depicted. ****p<0.0001 (Kruskall-Wallis test and Dunn’s multiple comparisons test). **b** Schematic of principle and treatment protocols of the micronucleus (MN) assays depicted in c and d. Cytokinesis is blocked with cytochalasin B (CytoB), allowing scoring of MN in binucleate (BN) cells. **c, d** Using the G0 and S MN assay, micronuclei were scored in untreated PHA blasts (c) and in PHA blasts exposed to 1 Gy of ionizing radiation (IR) (d). The Mitomycin C (MMC) MN assay was used to quantify micronuclei in PHA blasts treated with 0.02 µg/ml MMC (d). Mean MN values of a HC group are indicated by dashed lines. Dotted lines correspond to the mean+3SD and serve as a cut-off for sensitivity. Data of two independent experiments is shown for F1Pt, in each experiment an internal control (IC) was included. One experiment is performed for the family members (F1Si, F1Fa, F1Mo). **e** Bar graph depicts the total percentage of cell death after 120h of culture for untreated fibroblasts of HC, F1Pt, and F02-98. Mean and SD of at least three independent experiments is shown. **f** Schematic outlining of the treatment protocol of the cytotoxicity assay used in g and h. **g, h** Fibroblasts from HC, F1Pt, and F02-98 were untreated or exposed to genotoxic inducers (0.2 µg/ml MMC, 20 J/m² UV, or 10 Gy IR) without (g) and with (h) kinase inhibitors (20 nM ATRi or 10 µM ATMi) as indicated. Cell death was monitored by live imaging up to 120h by quantifying the number of cells stained with SYTOX Green. Cell death kinetics are reflective of at least two independent experiments. **i** Bar graph indicating % cell death after 120h of treatment for the experiments shown in g and h. Source data are provided as a Source Data file.

Next, we assessed chromosomal sensitivity to IR and MMC. No increase in radiation-induced MN yields was observed in F1Pt PHA blasts with the G0 MN assay, indicating a proficient NHEJ DSB repair pathway. Irradiation during the S phase led to a higher yield of MN (Fig. 7d and Supplementary Fig. 7b), highlighting a specific S phase sensitivity to radiation in absence of ATRIP, a phenotype not earlier associated with cells deficient in ATR signaling^12,61^. As anticipated, F1Pt cells also showed impaired ICL repair, reflected by the increase in MN formation in the presence of MMC (Fig. 7d). No increase in MN levels was detected in PHA blasts of the parents using both the MMC and S MN assay (Fig. 7c, d and Supplementary Fig. 7b), indicating that the heterozygous presence of the c.829+5G>T variant does not compromise ATR signaling.

Notably, persistent DNA damage accumulated during unperturbed replication in F1Pt and F02-98 fibroblasts negatively affected cell survival (Fig. 7e, g). In addition, we meticulously quantified cell death and cell proliferation in absence of functional ATR signaling following DNA damage induction by MMC, UV, or IR (Fig. 7f-i and Supplementary Fig. 7e, f). Treatment with 0.2 µg/ml MMC and 20 J/m² UV revealed a profound sensitivity of F1Pt and F02-98 fibroblasts, whereas cell survival of control fibroblasts was slightly impaired by MMC and unaffected by UV (Fig. 7g). These results confirm the previously described MMC and UV hypersensitivity in absence of functional ATR signaling^12,15,61^. In line with the G0 and S MN assay results of PHA blasts, exposure to 10 Gy IR only elicited a slight increase in cell death in F1Pt and F02-98 fibroblasts (Fig. 7g). Combined treatment of ATRi and a DNA damage inducer progressively compromised cell survival. Although minor in the case of F1Pt and F02-98 fibroblasts, this increase was consistently observed for all three genotoxic treatment conditions, suggesting residual ATR kinase activity (Fig. 7h).

In contrast to F1Pt and F02-98 fibroblasts, the combined treatment of ATRi with genotoxic stressors profoundly impaired cell survival in HC fibroblasts (Fig. 7h). To assess redundancy with the other DDR kinases, we quantified cell death rates in the presence of specific inhibitors targeting ATM and DNA-PKcs. ATM inhibition (ATMi) resulted in particularly high lethality in combination with ATRIP deficiency (F1Pt) or hypomorphic *ATR* variants (F02-98), contrasting with the response observed in HC fibroblasts (Fig. 7h and Supplementary Fig 7d). This was most evident upon IR-induced genotoxic stress, implying a substantial redundancy between the ATR and ATM kinase. Concomitant DNA-PK inhibition with genotoxic treatment did not evoke additional sensitivities in F1Pt, F02-98, or HC fibroblasts (Supplementary Fig. 7c, d). In conclusion, these findings demonstrate that loss of ATRIP results in chromosomal sensitivity and compromised cell viability, although some residual ATR signaling was retained. Moreover, concurrent ATM kinase activity is needed to mitigate the most detrimental effects of genotoxic stressors, as previously suggested for ATR deficient cells^15,61^.

### Modeling of ATRIP variants by CRISPR-Select^TIME^ exposes reduced cell fitness

Finally, to confirm the impact of the c.829+5G>T and c.829+2T>G *ATRIP* variants in an independent cellular model, cell fitness was assessed using the CRISPR-Select^TIME^ methodology^38^. To accurately study the effects of these variants, an MCF10A cell line harboring only one functional *ATRIP* or *ATR* allele was generated (Fig. 8a and Supplementary Fig. 8a). In addition to the variants identified in this study, a hypomorphic leaky splice variant in *ATR* (c.2022A>G, homozygous in F02-98^12^), the proposed causal *ATRIP* variants identified in patient CV1720^14^ (c.2278C>T and c.248-14A<G), and a homozygous benign *ATRIP* splice variant present in gnomAD v4.0.0 (c.2056-6_2056-3del) were modeled. As expected, the presence of the c.2022A>G *ATR* variant resulted in strongly reduced cell fitness (Fig. 8b). CRISPR-Select^TIME^ revealed similarly large effect sizes for both *ATRIP* splice variants, c.829+5G>T (homozygous in F1Pt) and c.829+2T>G (homozygous in F46.1). Moreover, we confirmed the anticipated neutrality of the homozygous *ATRIP* variant reported in gnomAD v4.0.0 (c.2056-6_2056-3del). Surprisingly, for the reported *ATRIP* variants of patient CV1720, reduced fitness was only evident for the c.2278C>T variant. Although abnormal splicing was observed in presence of the second *ATRIP* allele of patient CV1720, we were unable to validate the causality of the candidate variant c.248-14A>G using CRISPR-Select^TIME^ (Fig. 8c and Supplementary Fig. 8b), which might reflect the originally reported hypomorphic nature of this variant^14^. Given the role of both ATR and ATRIP in regulating DNA replication and proliferation dynamics, application of the CRISPR-Select^TIME^ methodology in a mono-allelic ATRIP cell line appeared particularly suitable to establish the pathogenic role of the identified *ATRIP* variants c.829+5G>T and c.829+2T>G, next to the known c.2278C>T variant. Moreover, frequencies of these variants and their corresponding internal controls (frameshifts) were strikingly comparable, indicating similar selection against both editing outcomes (Fig. 8c and Supplementary Fig. 8b).

**Figure 8.**
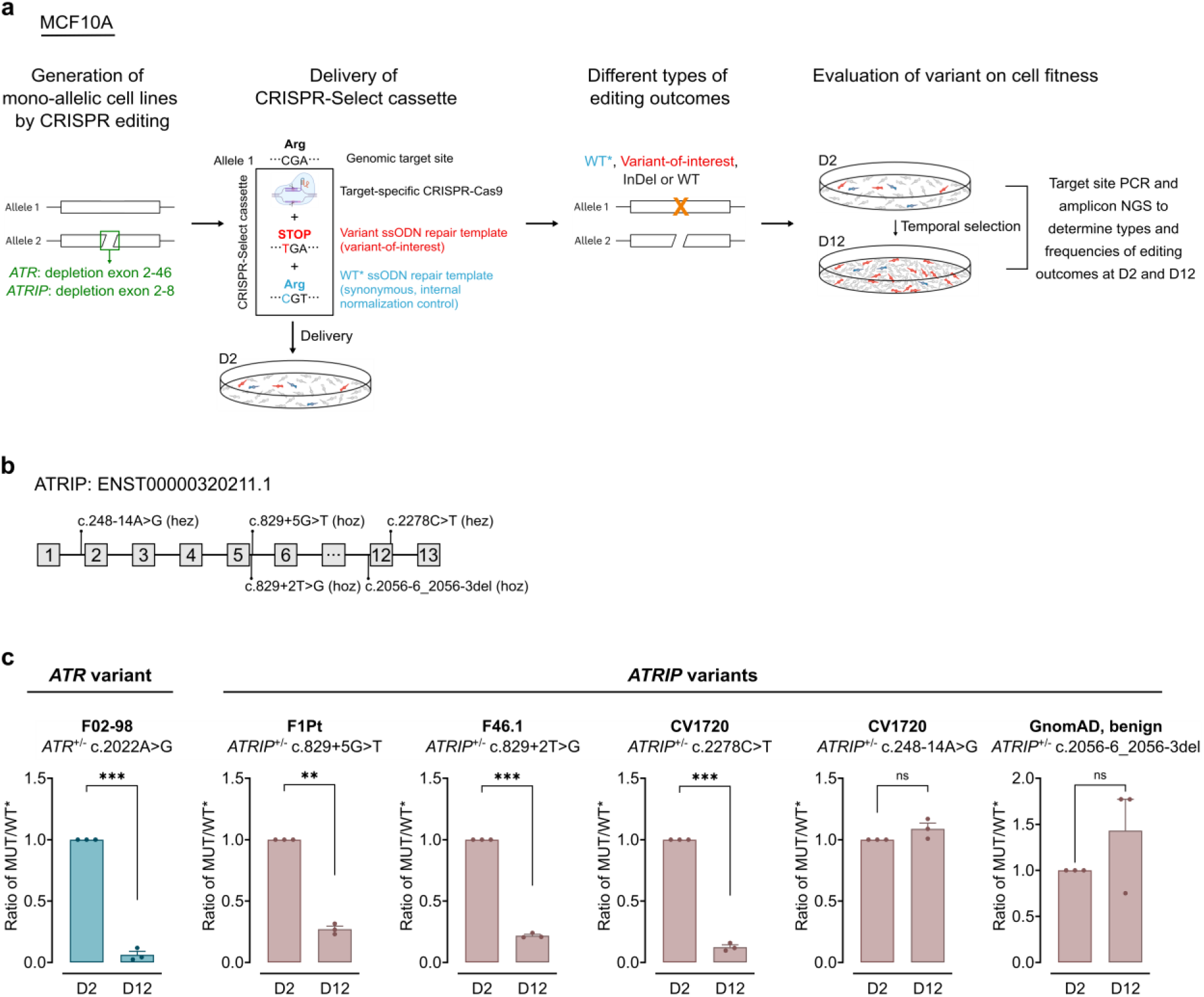
Quantitative functional assay identifies fitness defect in genome engineered *ATRIP* c.829+5G>T and *ATRIP* c.829+2T>G cells. **a** Schematic representation of CRISPR-Select^TIME^ process for cell fitness testing. The depletion region of the *ATR* and *ATRIP* genes are indicated. The CRISPR-Select cassettes for all the variants were transfected on day 2. Yellow cross indicates the various editing outcomes on target gene. The cell samples from day 2 (D2) and day 12 (D12) were collected and lysed for DNA extraction. PCR amplified the target edited sites of each sample and products were analyzed by NGS sequencing. **b** Schematic illustration of the modeled compound heterozygous (hez) and homozygous (hoz) *ATRIP* variants at the transcript level. **c** Cell fitness of MCF10A mono-allelic cells with the *ATR* and *ATRIP* variants. Each symbol within the bar represents an independent experiment. Each variant has three independent biological replicates. Mean and SEM are shown. Data of D12 is normalized to D2. **p<0.01, ***p<0.0001, ns: non-significant (two-tailed paired t-tests). Source data are provided as a Source Data file.

## DISCUSSION

In this study, we demonstrated that biallelic loss-of-function (LOF) *ATRIP* variants result in human disease, characterized by short stature, microcephaly, intellectual disability, and immunodeficiency. The causal role of the pathogenic *ATRIP* variants is supported by multiple lines of evidence; (1) the presence of homozygous novel or rare variants with a similar LOF consequence on protein level in two patients of independent ancestry, (2) the complete absence of homozygous LOF *ATRIP* variants in the non-affected population (cfr. gnomAD), (3) the finding of an impaired replication stress response associated with disturbed proliferation and survival in primary patient cells, and (4) reduced cell fitness when introducing these variants in an independent cellular model.

Ablation of ATR, TOPBP1, or CHK1 in murine models has consistently resulted in embryonic lethality, emphasizing the essential nature of the ATR signaling pathway^7,8,59,62,63^. This is further illustrated by the identification of exclusively hypomorphic *ATR* mutations in patients with microcephalic primordial dwarfism (MPD)^12,14–16^. In a similar vein, it has been suggested that ATRIP deficiency would be incompatible with life. However, our study demonstrates that LOF *ATRIP* variants are tolerated to some extent in human individuals, albeit resulting in a severe MPD phenotype. In line with our findings, knock-out *atrip* zebrafish were also shown to be viable but displayed a severely compromised body length and lifespan, suggesting a crucial role of *atrip* in growth and development during the juvenile stage^64^.

ATRIP plays a crucial role in facilitating stabilization, recruitment, and activation of the ATR kinase^11,40,65^. Studies using both cellular models and patient cells have shown that partial or complete ATRIP loss results in a significant reduction in ATR protein levels^11,14,41^. This co-dependency between ATR and ATRIP for protein stability is further evidenced by our data, showing markedly diminished ATR levels in complete absence of ATRIP. Several lines of evidence have demonstrated that ATRIP acts as a sensor of RPA-ssDNA nucleoprotein filaments, promoting ATR recruitment and subsequent activation of downstream substrates^17,65^. Interestingly, an early mutational study detected efficient CHK1 phosphorylation in the presence of mutated ATRIP devoid of its RPA-binding domain^41^. Correspondingly, our findings revealed that the remaining pool of ATR was able to form foci and autophosphorylate (T1989 phosphorylation) upon exposure to genotoxic stressors in absence of ATRIP, although at lower levels. These results suggest that ATRIP is required for ATR stability but not for ATR recruitment and activation. While both TOPBP1 and ETAA1 are able to activate ATR, only the ATR-TOPBP1 axis requires ATRIP for recruitment to RPA-ssDNA filaments^17,22,40^. It remains to be clarified whether binding and activation of ATR by ETAA1 is retained upon ATRIP loss. We speculate that interactions with ETAA1 and potentially other unanticipated complex partners may underly the ability of ATR to recognize DNA damage and display kinase activity in an ATRIP-independent manner.

Studies using murine models have provided compelling evidence implicating ATR signaling in the development, maturation, and maintenance of the immune system^66–70^, an association which had not been clearly described in humans to date. These mice consistently presented with developmental abnormalities analogous to MPD and features of premature ageing, demonstrating that unresolved replicative stress, proliferative failure, and cellular senescence likely underly both the developmental and immunological phenotype. The association of MPD with immunodeficiency is further supported by an emerging class of inborn errors of immunity (IEI) linked to monogenic defects in DNA replication factors^29,31–34,71^. These disorders typically present a combination of immunodeficiency with perturbed growth, microcephaly, progeroid features, and developmental abnormalities. All three ATRIP patients had clinical features suggestive for immunodeficiency, and the immunophenotype - characterized by lymphopenia and neutropenia - draws intriguing parallels to observations in other DNA replication-associated disorders. Our data revealed that ATRIP loss compromises the replication stress response, leading to a DNA repair signature in immune cells *in vivo* and impaired lymphocyte proliferation *in vitro*. Despite our observations in ATRIP deficient patients, no definite immunological phenotype has been observed in patients with biallelic *ATR* mutations (based on personal correspondence with respective clinicians). Although Mokrani-Benhelli et al.^15^ noted minor immune irregularities in a 9-year old patient with biallelic *ATR* mutations, drawing parallels to our patient F1Pt, the phenotype was relatively mild and no up-to-date immunological data could be obtained. Several explanations for these disparate observations are possible; (1) the hypomorphic nature of the *ATR* mutations may mitigate the impact of ATR deficiency on the human immune system, (2) ATR signaling in immune cells such as lymphocytes is critically dependent on ATRIP, (3) ATRIP has ATR-independent functions in the human immune system. In line with this, considerable heterogeneity in immunological features is also observed in both DNA repair and DNA replication-associated IEI, highlighting that mutations within the same cellular processes can lead to diverse phenotypes^31,32,71,72^. Finally, several older studies reported immunological abnormalities, such as pancytopenia, in patients with a phenotypical diagnosis of Seckel syndrome^73–76^. It is worth noting, however, that the molecular causes of Seckel syndrome had not yet been identified at that time, potentially leading to misdiagnoses due to overlapping features with other MPD-like disorders^77,78^. More studies, comprehensively describing the immunological phenotype in molecularly characterized disorders of DNA replication and DNA repair, will be required to further our understanding of the role of ATR and ATRIP in the biology of immune cells.

In summary, our study provides evidence supporting *ATRIP* as a disease-associated gene in MPD and IEI and offers valuable insights into the cellular and immunological characteristics. Our findings challenge the conventional paradigm of the ATR-ATRIP interaction, underscoring the need for further investigation into the molecular mechanisms governing ATR activation upon DNA damage. Although our data suggests an essential role for ATRIP in safeguarding immune cells against the consequences of unresolved replicative stress, it remains unclear whether this reflects an unidentified noncanonical function of ATRIP or this is mediated by the ATR signaling pathway. Careful clinical monitoring and additional research are needed to further delineate the phenotypic spectrum associated with both biallelic *ATR* and *ATRIP* mutations and to better understand the cellular mechanisms driving the immunological phenotype.

## MATERIAL AND METHODS

### Study approval

Ethical approval for this study was granted by the ethics committee of Ghent University Hospital in Belgium (2012/593 and 2019/1565). Clinical data and samples were collected with informed consent from the participants of the study, in accordance with the 1975 Helsinki Declaration.

### Cell culture

Lymphoblastoid cell lines (LCLs) were derived from blood following EBV transformation. LCLs were maintained in Roswell Park Memorial Institute (RPMI) 1640 medium supplemented with GlutaMAX, 10% FBS (Bodinco and Tico Europe), 1% penicillin-streptomycin (10,000 U/mL; Gibco), 1 mM sodium pyruvate (Gibco) and 50 μM 2-mercaptoethanol (Gibco). PHA blasts were generated by culturing cryopreserved isolated peripheral blood mononuclear cells (PBMCs) in LCL medium supplemented with 2% phytohemagglutinin-M (PHA-M; Gibco). HEK293T cells (The American Type Culture Collection (ATCC)) were cultured in Dulbecco’s modified Eagle’s medium (DMEM) supplemented with 10% FBS, 1% penicillin-streptomycin (10,000 U/ml), 2mM GlutaMAX (Gibco). Primary human dermal fibroblast cells were cultured in DMEM supplemented with 10% FBS, 1% penicillin-streptomycin (10,000 U/ml), 2mM GlutaMAX, 0.1 mM sodium pyruvate, and 50 µM 2-mercaptoethanol. MCF10A cells (ATCC; CRL-10317) were cultured in Dulbecco’s Modified Eagle Medium:Nutrient Mixture F-12 (DMEM / F-12) (Thermo Fisher Scientific; 31330038), supplemented with 5% horse serum (Thermo Fisher Scientific; 16050122), 1% Penicillin/Streptomycin (Thermo Fisher Scientific; 15070063), 10 μg/ml insulin (Sigma-Aldrich; I1882), 0.5 μg/ml hydrocortisone (Sigma-Aldrich; H0888-1G), 20 ng/ml EGF (Peprotech; AF-100-15), and 100 ng/ml cholera toxin (Sigma-Aldrich; C8052-5MG). ATR deficient fibroblasts from a previously described ATR patient (F02-98) were purchased from Coriell institute (GM18366)^12^. All cell lines were incubated at 37°C with 5% CO2.

### Sequencing analysis

Trio whole exome sequencing (WES) on genomic DNA of the patient and unaffected parents was performed using KAPA HyperExome probes (Roche). Paired-end massive parallel sequencing was performed on a NovaSeq 6000 Instrument (Illumina). Data analysis was performed on an in-house developed platform, in accordance with the American College of Medical Genetics and Genomics and the Association for Molecular Pathology (ACMG-AMP) guidelines^79^. Variant confirmation and segregation analysis was performed by Sanger Sequencing on an ABI 3730 platform (Applied Biosystems).

RNA for evaluation of aberrant splicing was obtained using the Maxwell® RSC simplyRNA Tissue Kit on a Maxwell® RSC Instrument (Promega Corporation) from PHA blasts stimulated with 1 μl/ml PHA-M and 50 μl/ml interleukin-2 (IL-2) (Sigma-Aldrich), LCLs, and patient-derived dermal fibroblasts. cDNA was synthesized using Superscript III Reverse Transcriptase (Invitrogen) according to manufacturer’s recommendations. Amplicon length analysis was performed using a TapeStation (Agilent Technologies) or an ABI 3730 platform (Applied Biosystems).

Targeted RNA-sequencing was performed with the SureSelectXT HS2 RNA System kit (Agilent Technologies) using SureSelectXT Human All Exon V7 probes (Agilent Technologies) for target enrichment and sequencing was performed on a NovaSeq 6000 Instrument (Illumina).

All primers were purchased from Integrated DNA Technologies and are listed in Supplementary Table 5. Nucleotide and protein numbering is in accordance with transcript ENST00000320211 and NM_000057.3, respectively.

### RT-qPCR

Fibroblasts from healthy controls and patient F1Pt were lysed in RLT buffer (Qiagen) and stored at - 80°C until further processing. RNA was purified using the RNeasy Mini kit (Qiagen) according to manufacturer’s protocol. Total RNA was reverse transcribed into cDNA using the SensiFast cDNA Synthesis Kit (Bioline) and real-time quantitative PCR (RT-qPCR) was performed using the LightCycler 480 (Roche). Gene transcript levels were normalized against an endogenous B-actin control. Samples were run in duplicates in two independent experiments. Results are depicted as the relative expression compared to the healthy controls, determined using the 2^−ΔΔCt^ method. To determine mRNA expression levels of a shorter mutant transcript and a transcript containing exon 5 in F1Pt fibroblasts, primers against an amplicon in exon 3-4 and exon 5-6 of ATRIP were used, respectively. Predesigned qPCR primer pairs were ordered from Integrated DNA technologies (https://eu.idtdna.com) (primers are listed in Supplementary Table 5).

### DNA damage inducers and inhibitors

Cells were irradiated with 254nm ultraviolet radiation (UV-C) in the absence of culture medium using the UVP Crosslinker (Analytikjena) or with X-rays (220kV, 13mA, 0.15mm copper filter, 3Gy/min) using the Small Animal Radiation Research Platform (SARRP) (Xstrahl) at indicated doses. The following genotoxic inducers and chemical inhibitors were used: Mitomycin C (MMC; Sigma-Aldrich), Hydroxyurea (HU; Sigma-Aldrich), ATR kinase inhibitor (ATRi) (20 nM; BAY-1895344, also known as Elimusertib), ATM kinase inhibitor (ATMi) (10 µM; KU-55933) and DNA-PK inhibitor (DNA-PKi) (2 µM; KU-57788, also known as NU7441). Inhibitors were added to the medium 1h prior to treatment with genotoxic stressors.

### Plasmids and cloning

Plasmid DNA constructs containing the wild type cDNA sequence for ATRIP, ATR, and RPA1 inserted into a modified pcDNA3.1(+) expression vector encoding a N-terminal 3xFlag-tag, a C-terminal V5-tag, or a C-terminal c-Myc-tag, respectively, were purchased from GenScript. Mutant ATRIP plasmid constructs containing the cDNA sequence for wild type ATRIP lacking exon 5 inserted into a pcDNA3.1(+) vector encoding a N-terminal 3xFlag-tag were generated by GenScript. All plasmids were transformed in DH10B or DH5α Escherichia coli. DNA was isolated using NucleoBond Xtra Midi kit (Macherey-Nagel). HEK293T cells were transiently co-transfected with ATR, RPA1, and either wild type ATRIP or mutant ATRIP plasmid DNA. Transfections were performed with branched 25kDa polethylenimine (PEI).

### Co-immunoprecipitation

Wild type or mutant ATRIP protein was immunoprecipitated by incubating 250 µg of whole cell lysate with 4 µg of anti-FLAG antibody (Sigma-Aldrich; F3165) or 4 µg of isotype-matched IgG antibody (Sigma-Aldrich; 12-371) overnight at 4°C, followed by incubation with Dynabeads Protein G (Invitrogen) for 1h at 4°C. Beads with protein complexes were washed six times with IP High buffer without extra salt and detergents (Nuclear complex Co-IP Kit; Active Motif; 54001). Precipitates were eluted by boiling in Laemmli Sample buffer and analyzed by immunoblotting.

### Immunoblotting

Cells were lysed in RIPA (150 mM NaCl, 1.0% IGEPAL CA-630, 0.5% sodium deoxycholate, 0.1% SDS, 50 mM Tris, pH 8.0) (Roche) complemented with protease inhibitors (cOmplete ULTRA; Roche) and phosphatase inhibitors (PhosSTOP; Roche). Soluble fraction was normalized prior to protein separation by SDS-PAGE using a 4–15% Criterion TGX Stain-Free Protein Gel (Bio-Rad) and followed by semi-dry transfer to a nitrocellulose membrane (Bio-Rad). Membranes were blocked with 5% nonfat dry milk (Cell Signaling Technology (CST)) in phosphate buffered saline with 0.1% Tween 20 (PBS-T) or 5% bovine serum albumin (BSA) (Roche) in PBS-T for 1h at room temperature, incubated with primary antibodies overnight at 4°C or for 2h at RT, followed by incubation with secondary antibodies conjugated to horseradish peroxidase (HRP). Membranes were developed using enhanced chemiluminescence reagents (SuperSignal West Dura or Femto; Thermo Fisher Scientific) on a Chemidoc imaging system (Bio-Rad). All immunoblotting experiments were performed at least twice, and representative data are shown. Protein levels were quantified using the Image Lab software (Bio-Rad). All uncropped images can be retrieved in the Source File. The following primary antibodies were probed: anti-Flag (1:1000; Sigma-Aldrich; F3165), anti-V5 (1:1000; Invitrogen; R960-25), anti-c-Myc (1:2000; Abcam; ab9106), anti-ATRIP (1:1000; Cell Signaling Technology (CST); #97687), anti-TOPBP1 (1:2000; Bethyl Laboratories; A300-111A), anti-RPA70/RPA1 (1:2000; Abcam; ab176467), anti-ATR (1:1000; CST; #2790), anti-CHK1 (1:1000; Santa Cruz; sc-8408), anti-phospho-ATR (T1989) (1:1000; Invitrogen; PA5-77873), anti-phospho-CHK1 (S317) (1:1000; CST; #8191), anti–β-tubulin– HRP (1:5000; Abcam; ab21058) and anti-GAPDH-HRP (1:2000; CST; #8884). The secondary antibodies goat anti-mouse IgG (H+L)-HRP (1:10000; Invitrogen; G-21040) and goat anti-rabbit IgG-HRP (1:2000; CST; #7074) were used. To confirm the specificity of the anti-phospho-ATR antibody, the membrane was dephosphorylated using Lambda Protein Phosphatase (New England Biolabs; P0753S). Briefly, the membrane was incubated with blocking buffer containing 400 U/ml Lambda Protein Phosphatase and 1 mM MnCl2 overnight at 4°C prior to incubation with the primary antibody.

### Immunofluorescence

Primary fibroblasts were grown on 22 mm glass coverslips (Menzel Gläser) 24h before starting the experiment. Cells were pulse-labeled with 10 µM EdU (Click-iT™ Plus EdU Alexa Fluor™ 647 Imaging Kit; Invitrogen; C10640) for 30 min and concomitantly treated with a DNA damage inducer. For RPA staining, cells were treated with pre-extraction buffer (0.5% Triton X-100, 20 mM HEPES, pH 7.4, 100 mM NaCl, 3 mM MgCl2, and 300 mM sucrose in distilled water) prior to fixation. At the indicated time post-treatment, cells were fixed with 3% paraformaldehyde (PFA) in PBS for 20 min, followed by simultaneous permeabilization and blocking in 1% BSA (Sigma-Aldrich) and 0.2% Triton X-100 in PBS for 30 min. Primary antibody incubations were performed for 1h at RT with anti-γH2AX S139 (1:500; BioLegend; 613402) and anti-RPA32/RPA2 (1:500; Merck; MABE285) or overnight at 4°C with anti-ATR (1:250; CST; #13934). After washing, coverslips were incubated with secondary antibodies for 1h at RT (goat anti-rabbit Alexa Fluor 488 (1:1000; Invitrogen; A-11008) or goat anti-mouse Alexa Fluor 488 (1:1000; Invitrogen; A-11001)). EdU staining was performed by incubation with Click-iT reaction cocktail according to the manufacturer’s protocol. Coverslips were mounted in Fluoromount (Sigma-Aldrich) containing DAPI nuclear stain. Images were acquired with a Leica DM6 B microscope (Leica Microsystems) using a HCX PL APO 40x/1.3 oil objective, equipped with Las X software (Leica Microsystems). Identical microscope settings and image processing steps were used for all samples of the same experiment. Quantifications of DNA damage foci and analysis of signal intensities was performed using Fiji software (version 2.9.0). Pooled data from three independent experiments are shown. A minimum of 150 cells were analyzed per experimental condition.

### Micronucleus assays

#### The G0 and MMC micronucleus assay

The G0 and MMC micronucleus (MN) assay were performed, as described previously by Beyls et al.^80^ and Francies et al.^81^, respectively. Briefly, fresh whole blood cultures were prepared by adding 0.5 ml blood to 4.5 ml RPMI. The culture medium was supplemented with 10% FBS and 1% penicillin-streptomycin. The cultures were irradiated with 0.5 or 1 Gy (G0 MN assay) or treated with 0.02 µg/ml MMC (MMC MN assay) and subsequently stimulated with 2% PHA-M. Cytochalasin B (6 µg/ml; Sigma-Aldrich) was added to block cytokinesis 23h post-stimulation. After 70h culture time, cells were harvested and exposed to a cold hypotonic shock with 0.075 M KCl and fixed in ice-cold methanol/acetic acid solution. Acridine orange (10 µg/ml; Sigma-Aldrich) stained slides were scored according to the criteria described by Fenech et al.^82^ MN were scored manually on a fluorescence microscope in 1000 binucleate (BN) cells per condition.

#### The S micronucleus assay

Fresh whole blood cultures were set-up as described above and cell proliferation was immediately stimulated with 2% PHA-M. Following 96h of culture, the cells were pulse labeled with 10 µM EdU for 30 min, irradiated with 0.5 or 1 Gy, and cytochalasin B (6 µg/ml; Sigma-Aldrich) was added. After an additional 24h of culture, the cells were harvested and fixed according to the G0 MN protocol described previously. Slides were prepared and the cells were additionally fixed in 3% PFA for 20 min and permeabilized with ice-cold 0.2% Triton X-100 for 10 min. Subsequently, EdU staining was performed according to the manufacturer’s protocol and the slides were mounted in Fluoromount containing DAPI nuclear stain. MN were scored in 1000 EdU positive BN cells per condition.

### Flow cytometry

#### Cell cycle analysis

Fibroblasts were seeded at a density of 7,000 cells/cm² and cultured for 24h. The cells were exposed to a DNA damage inducer (0.02 µg/ml MMC, 200 J/m² UV-C, or 1 mM HU) and subsequently cultured for the indicated time. The cells were pulse-labeled with 10 µM EdU for 30 min before harvesting. The cells were fixed and permeabilized using the Foxp3/Transcription Factor Staining Buffer Set (Thermo Fisher Scientific; 00-5523-00) following the manufacturer’s protocol. EdU staining was performed by incubating the cells with the Click-iT reaction cocktail, prepared as per manufacturer’s instructions. The cells were stained with permeabilization buffer containing anti-γH2AX-AF488 (S139) (2F3; BioLegend; 613406) for 30 min on ice when indicated. DNA content was stained using permeabilization buffer containing DAPI. Acquisition of the stained cells was performed using a BD LSRFortessa or BD FACSymphony A3 (BD Biosciences). Subsequent data analysis was performed with the FlowJo v10.10.0 software (BD Life Sciences).

#### EdU pulse-chase assay

The EdU pulse-chase assay was performed as previously described by Duthoo et al.^60^, with minor adjustments. Briefly, PHA blasts were cultured for 96h in a 48-well plate (250,000 cells in 500 µl) in the presence of 2% PHA-M. Next, the cells were pulse-labeled with 10 µM EdU for 30 min and concurrently exposed to a DNA damage inducer (200 J/m² UV-C or 4 Gy IR). MMC (0.02 µg/ml) was added to the culture 24h prior to EdU labeling. Subsequent harvesting of the cells was performed at various time points, ranging from 0 to 15h, with intervals of 3h. The cells were fixed and permeabilized using the Foxp3 / Transcription Factor Staining Buffer Set. Subsequent EdU staining was performed using the Click-iT reaction cocktail and DNA was stained with permeabilization buffer containing DAPI. Acquisition was performed using a BD LSRFortessa (BD Biosciences) and data was analyzed with the FlowJo software.

#### Proliferation assay

Cryopreserved PBMCs were resuspended in PBS at a density of 1 × 10^6^ cells per ml. Cells were labeled with CellTrace Violet (CTV) (1 µM; Thermo Fisher Scientific; C34557) and incubated for 10 min at 37°C. Unbound CTV was quenched by washing the cells with PBS supplemented with 1% FCS. The cells were seeded in a 48-well plate (250,000 cells in 500 µl), treated with 0.02 µg/ml MMC, and subsequently stimulated with 2% PHA-M. After 96h of culture, PHA blasts were harvested and stained with anti-CD3-AF700 (SK7; BioLegend; 344822), anti-CD4-FITC (RPA-T4; BD Bioscience; 561005), anti-CD8-PE-Cy7 (RPA-T8; BD Bioscience; 557746), and FcR block (BioLegend; 422302) in PBS. After 30 min of staining, cells were washed, stained with propidium iodide (PI), and acquired using a BD LSRFortessa (BD Biosciences). Subsequent data analysis was performed with the FlowJo software.

#### Immunophenotyping

Cryopreserved PBMCs were thawed in 37°C preheated complete medium (RPMI-1640 medium supplemented with GlutaMAX, 10% FCS, 1% penicillin-streptomycin (Pen/Strep) (10,000 U/mL; Gibco; 15140122), 1 mM sodium pyruvate (Gibco; 11360070), 1% non-essential amino acids (NEAA) (Gibco; 11140035), and 50 μM 2-mercaptoethanol (Gibco; 31350010)). Cells were left to recuperate for 30 min at 37°C and 5% CO2 after removal of DMSO. Cells were counted and 2,000,000 cells were plated for each panel. Cells were first stained with FcR block (BioLegend; 422302) together with biotin conjugated antibodies and Zombie UV™ Fixable Viability dye (BioLegend; 423107) in PBS. In a second step, a first set of surface markers were stained with a mixture of antibodies in FACS buffer (DPBS pH7.4, 1% Bovine Serum Albumin, 0.05% NaN3, 1 mM EDTA) and Brilliant Stain buffer (BD Biosciences). After 30 min of staining, cells were washed and stained overnight with a second set of antibodies. Cells were fixed, permeabilized, and intracellular stained with antibodies using the Foxp3/Transcription Factor Staining Buffer Set according to the manufacturer’s protocol. Acquisition and analysis of labeled cell suspensions was performed with a FACSymphony flow cytometer (BD biosciences) and subsequent analysis of data with the FlowJo software. Antibodies used to define PBMC populations can be found in Supplementary Table 6.

### Single-cell RNA, TCR, BCR, and CITE sequencing

After thawing, up to 2×10^6^ PBMCs of each individual were counted, isolated, and spun down. The cell pellet was resuspended and incubated for 30 min on ice with staining mix in PBS containing 0.04% BSA, CD19-PE-Cy7, eFluor506 Fixable Viability dye, Human TruStain FcX (BioLegend; 422302), and TotalSeq-C hashing antibodies (1:500; BioLegend). Two patient samples were multiplexed per lane using TotalSeq-C Cell Hashing Antibodies. Both total live PBMCs and B cells were sorted and loaded on different lanes. Sorted single-cell suspensions were resuspended at an estimated final concentration of 2,000 cells/µl and loaded on a Chromium GemCode Single Cell Instrument (10x Genomics) to generate single-cell gel beads in emulsion. The scRNA/Feature Barcoding/TCR libraries were prepared using the GemCode Single Cell 5′ Gel Bead and Library kit, version 1.1 (10x Genomics; 1000165), according to the manufacturer’s instructions. The cDNA content of prefragmentation and post-sample index PCR samples was analyzed using the 2100 BioAnalyzer (Agilent). Sequencing libraries were loaded on an Illumina NovaSeq flow cell at Vlaams Instituut voor Biotechnologie (VIB) Nucleomics core with sequencing settings according to the recommendations of 10x Genomics, pooled in a 75:20:5 ratio for the gene expression, TCR, and antibody-derived libraries, respectively.

### LAM-HTGTS

To study CSR patterns in B cells from F1Pt and healthy controls, a modified version of the linear amplification-mediated high-throughput genome-wide translocation sequencing (LAM-HTGTS) method was used, as described in Takada et al.^49^.

### Cytotoxicity assay

Fibroblasts were seeded into 24-well plates at a density of 6,000 cells/cm² 24h before starting the experiment. Cells were exposed to a DNA damage inducer (0.2 µg/ml MMC, 20 J/m² UV-C or 10 Gy IR) and incubated with SYTOX Green nucleic acid stain (1:100000; Invitrogen) to indicate dead cells during cell culture. Brightfield and fluorescence images were captured for 5 days with an 8h interval with the CELLCYTE X (Cytena). Cell confluency and the number of dead cells were quantified using CELLCYTE X Analysis software.

### CRISPR-Select^TIME^ in mono-allelic cell system

#### Generation of mono-allelic ATR^+/−^ and ATRIP^+/−^ cell lines

An iCas9-MCF10A-WT clonal cell line with stably integrated TRE3G Edit-R Inducible Lentiviral Cas9 construct (Horizon; CAS11229) was gifted by Roderick L. Beijersbergen, The Netherlands Cancer Institute. To generate ATR^+/−^ cells, iCas9-MCF10A-WT cells were transfected with dual gRNAs targeting intron 1 and intron 46 of the *ATR* gene. ATRIP^+/−^ cells were generated by transfecting the iCas9-MCF10A-WT cells with dual gRNAs targeting intron 1 and intron 8 of the *ATRIP* gene. Two CRISPR RNAs (crRNAs) for Streptococcus pyogenes Cas9 were designed using the online software Crispor (http://crispor.tefor.net/crispor.py). Note, the crRNAs should be located in the non-coding region to avoid disturbing adjacent genes. The knock-out efficiency of the designed crRNAs was analyzed by the online tool ICE (https://ice.synthego.com/<u>#/</u>), and subsequently, the crRNAs with a high knock-out efficiency were selected to establish the mono-allelic cell line. The iCas9-MCF10A-WT cells were seeded with 60% confluency in a 6 cm culture dish. To induce the expression of the cas9 protein, 1 μg/ml doxycycline was added to the medium. After 24h, 7.5 µl of each crRNA (2 nmol) was mixed with 15 µl of trans-activating crRNA (tracrRNA, 5 nmol) and was incubated at room temperature for 10 min to form crRNA:tracrRNA duplexes. Cells were subsequently transfected by 4 ml of culture medium containing 30 µl crRNA:tracrRNA duplexes, 15 µl lipofectamine RNAiMAX (Thermo Fisher Scientific; 13778), and 500 µl Opti-MEM (Thermo Fisher Scientific; 31985062). After 2 days of culture, single cells were sorted into 96-well plates with 100 µl fresh medium per well using a FACS Aria III instrument (BD Biosciences) and subsequently kept in culture 2-3 weeks until clonal cell lines were formed. Genomic PCR was performed to evaluate the mono-allelic cell line. Primers targetting WT, *ATR,* or *ATRIP* deletion alleles were designed through Primer3 (https://primer3.ut.ee/). Primers annealing to sequences outside the depletion region were used to identify the knock-out alleles. Primers annealing to sequences inside the deleted regions were used to detect the WT allele. The cell colonies containing DNA that is successfully amplified by both two types of primers were mono-allelic cell lines. DNA extraction was performed using the Quick-DNA^TM^ Miniprep Plus Kit (Zymo Research; D4069). PCR was performed using 100 ng genomic DNA and 1% agarose gel electrophoreses. For further verification, the PCR products of target sites were purified using the QIAquick Gel Extraction Kit (QIAGEN; 28706) and were sent to Eurofins for Sanger sequencing. The designed crRNAs and PCR primers are listed in Supplementary Table 7.

#### In vitro CRISPR-Select^TIME^ assay

In a 6-well plate, 17,500 iCas9-MCF10A-ATR^+/−^ or ATRIP^+/−^ cells were seeded in 2 ml culture medium per well. To induce the expression of Cas9 protein, 1 μg/ml doxycycline was added in each well 24h before cell transfection. After 24h, 7.5 µl of the designed crRNA (2 nmol) was incubated for 10 min at room temperature with 7.5 µl of tracrRNA (5nmol). Next, a mixture of 15 µl crRNA:tracrRNA duplexes, 7.5 µl lipofectamine RNAiMAX, 250 µl Opti-MEM, and 2 µl mixture of designed variant (10 µM) and WT* single-stranded oligodeoxynucleotide (ssODN) repair templates was dripped onto the iCas9-MCF10A cells in fresh medium. The DNA repair template for each variant was designed in the online software Benchling (https://benchling.com). The design of the synonymous mutations referring to the codon usage information were posted on https://www.kazusa.or.jp. After 2 days, 50% of the edited cells were collected and DNA extraction was performed using the Quick-DNA^TM^ Miniprep Plus Kit. The remaining edited cells were kept in culture and passaged every 3 days. After an additional 10 days of culture, all cells were collected and lysed for DNA extraction using the Quick-DNA^TM^ Miniprep Plus Kit. Using Benchling and Primer3, PCR primers for the amplification of the edited target sites in each variant were designed. Two rounds of PCR were performed to prepare the PCR products for subsequent amplicon NGS. Firstly, primer pairs were used to amplify the specific edited target sites, containing overhangs with binding sites for the primers of the second round. Secondly, specific barcodes were added to each sample, including overhangs with barcodes or adaptors for NGS. After purifying the PCR products using the QIAquick Gel Extraction Kit, NGS was performed to sequence the PCR products of the DNA samples collected on day 2 and day 12. The ratio of read numbers of the knock-in of a variant relative to the WT* on day 2 and day 12 were compared to determine the functional characteristics of the different mutations. The designed crRNAs, PCR primers, and DNA repair templates are listed in Supplementary Table 8 and 9.

### Statistical analyses

GraphPad Prism software (version 10) was used for all statistical analyses. As indicated in the figure legends, results were analyzed with a two-tailed Mann-Whitney test, multiple paired t-tests, or Kruskal-Wallis test combined with Dunn’s multiple comparisons test. The threshold for statistical significance was set to p<0.05. Representative immunofluorescence images and flow cytometric plots are shown. R (v4.3.2) and RStudio (v2023.09.1+494) were used for analysis of scRNA, TCR, BCR, and CITE sequencing data. The analytical pipeline consisted of Cellranger (v6.1.1) for quality control. CITE-seq and scRNA-seq data were analyzed using Seurat package (v5), with an embedded Presto package (v1.0.0) for differential gene expression analysis. Analysis of TCR-seq and BCR-seq data was conducted using the Diversity AnaLysis Interface (DALI, v2.0.0).

## Statements

Personally identifiable patient information was redacted in accordance with medRxiv requirements. IDs used in this study were not known to anyone outside the research group, unless they were previously published and are thus part of the public domain (PMID23144622 and PMID35568357).

## Acknowledgments

We gratefully thank the patients and family for consenting to this research. The valuable input from prof. Penny Jeggo, prof. Grant Stewart, prof. Mark O’Driscoll, Dr. Angela Brady, Dr. Mohnish Suri, Dr. Pradeep Vasudevan, Dr. Emma Hobson, Dr. Kaljit Bhuller, and Dr. Mandal Kausik on the clinical phenotype of the ATRIP and ATR patients is warmly appreciated. We thank Karlien Claes for the patient centered support, Sylvie De Buck for the administrative assistance, Lieselotte Vande Walle and Sarah Ghistelinck for the experimental expertise, and Julie Smet for providing NK cytotoxicity data. We acknowledge the VIB Flow Core and VIB Single Cell Core of the VIB center for inflammation research for their assistance and expertise in experimental design.

## Author contributions

Conceptualization: ED, EB, LB, TG, AB, AV, CSS, QPH, FH, KBMC, SJT

Methodology: ED, EB, LB, TG, AB, AV, CSS, QPH, FH, KBMC, SJT

Investigation: ED, EB, LB, PH, LJ, SR, BP, LD, VDB, MDB, JS, SJT

Clinical data collection: LH, TK, GM, FH

Visualization: ED, EB, LB, SJT

Funding acquisition: SR, EVB, AV, AB, FH, KBMC, SJ

Project administration: ED, EB, LB, SJT

Supervision: FH, KBMC, SJT

Writing – original draft: LB, ED, EB, SJT

Writing – review & editing: All authors

## Data availability

Code for scRNA-seq data analysis will be made available on GitHub upon publication. The plasmids will be made available via GeneCorner upon publication (https://bccm.belspo.be/genecorner). The *ATRIP* variant will be submitted to ClinVar and the Leiden Open Variation Database (LOVD) upon publication.

## Materials & Correspondence

Correspondence and requests for materials should be addressed to Filomeen Haerynck and Kathleen Claes.

