## Supplementary Materials for "ATRIP deficiency impairs the replication stress response and manifests as microcephalic primordial dwarfism and immunodeficiency"

**The file includes:**

Case description

Supplementary Figures 1-8

Supplementary Tables 1-9

### Case descriptions

*Personally identifiable patient information was redacted in accordance with medRxiv requirements.*

### Supplementary Figures

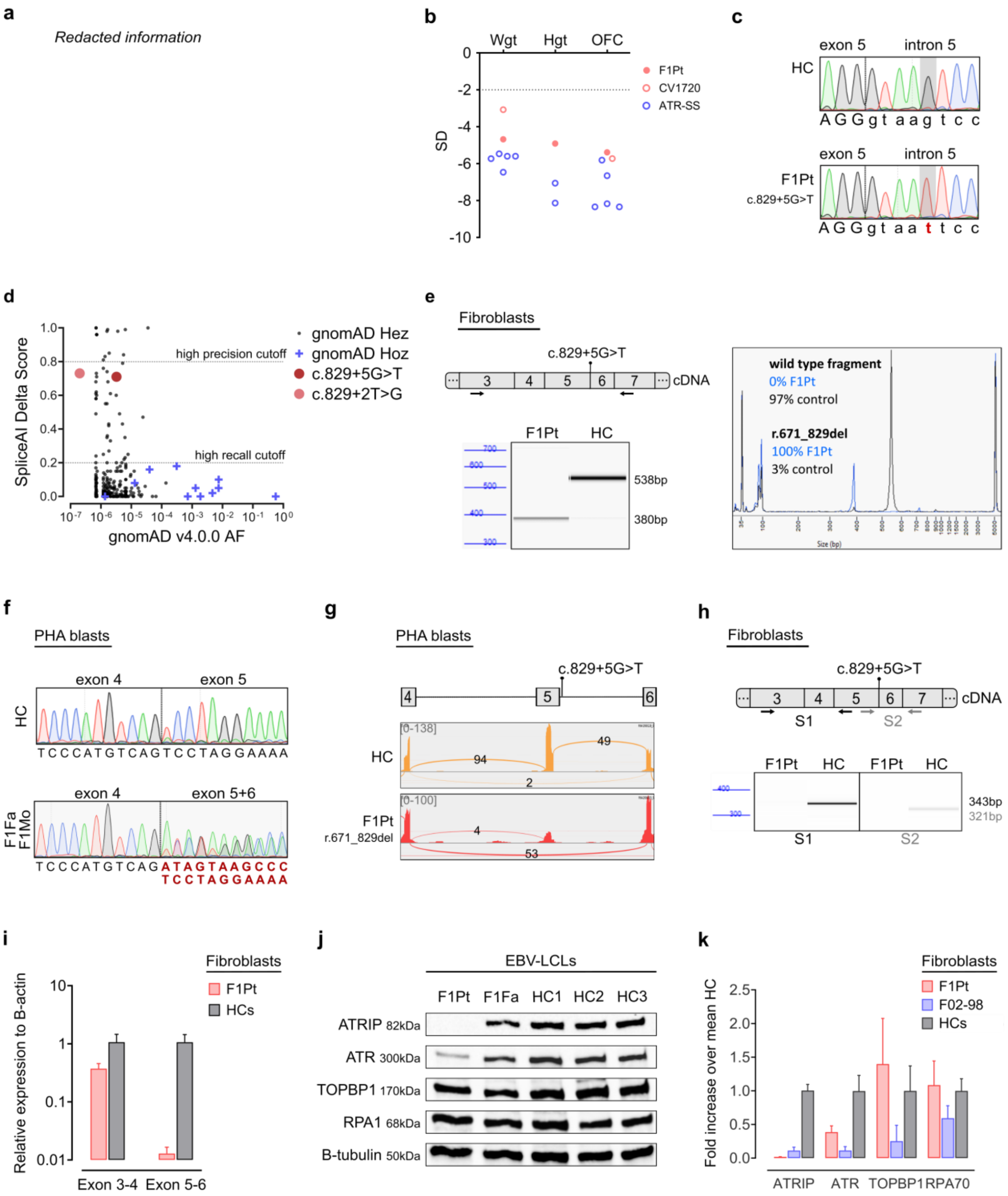


**Supplementary Figure 1.**

**a** Photographs of patients F1Pt and F46.1 demonstrating facial similarities, including sloping forehead and beak-like nose. **b** Weight (Wght), height (Hgt), and head circumference (occipital frontal circumference; OFC) at birth plotted as z-scores (SD from population mean for age and sex). Dashed line at -2SD indicates cut-off for normal population distribution. ATRIP patients are denoted by red dots, ATR patients are denoted by blue dots. **c** Electropherograms of genomic DNA extracted from blood for F1Pt and a healthy control (HC). Nucleotide numbering is in accordance with ENST00000320211.1. Images represent results from 5 independent experiments. **d** Population genetics: Highest SpliceAI Delta Score against gnomAD v4.0.0. allele frequency (AF) for splice region variants in *ATRIP* (ENST00000320211.1). Splice region variants are defined as nucleotide changes within the ±20 base pairs (bp) flanking the exon. Black dots and blue cross signs represent heterozygous and homozygous variants, respectively. More details regarding homozygous splice variants can be found in Supplementary Table 4. Red shaded dots represent *ATRIP* variants of interest (c.829+5G>T and c.829+2T>G). **e** Fragment analysis and size profiles of PCR-amplified cDNA extracted from fibroblasts for F1Pt and a HC. Arrows indicate the position of forward and reverse primers used for PCR amplification. Percentages represent relative quantification of the 538bp wild type and 380bp mutant (r.7671_829del) fragment. Data are reflective of 2 independent experiments. **f** Electropherograms of cDNA extracted from PHA blasts for F1Fa (father), F1Mo (mother), and a HC. Nucleotide numbering is in accordance with ENST00000320211.1. Data are reflective of five independent experiments. **g** Sashimi plot of targeted RNA-seq data generated in Integrative Genomics Viewer (IGV). Input RNA was extracted from PHA blasts of F1Pt and a HC. Exon numbering is in accordance with ENST00000320211.1. **h** Fragment analysis of PCR-amplified cDNA using two primer pairs (S1: E3-E5; S2: E5-E7, indicated by arrows) on fibroblasts from F1Pt and a HC. Data is reflective of two independent experiments. **i** Real-time quantitative PCR (RT-qPCR) analysis on fibroblasts of F1Pt and HCs (n = 3) of amplicon in exon 3-4 and exon 5-6. The relative expression to β-actin in a logarithmic scale is shown. Data from 2 independent experiments is shown. **j** Endogenous protein expression of ATRIP and interaction partners in EBV immortalized lymphoblastoid cell lines (EBV-LCLs) from F1Pt, F1Fa, and HCs (n = 3). B-tubulin was used as loading control. Western blot image is reflective of two independent experiments. **k** Quantification in arbitrary units of digitized chemiluminescent signals from Fig. 1E normalized to β-tubulin signal from the same lane. Graph depicts fold increase of normalized protein levels over the mean of HCs (n = 3) of 4 immunoblots. Source data are provided as a Source Data file.

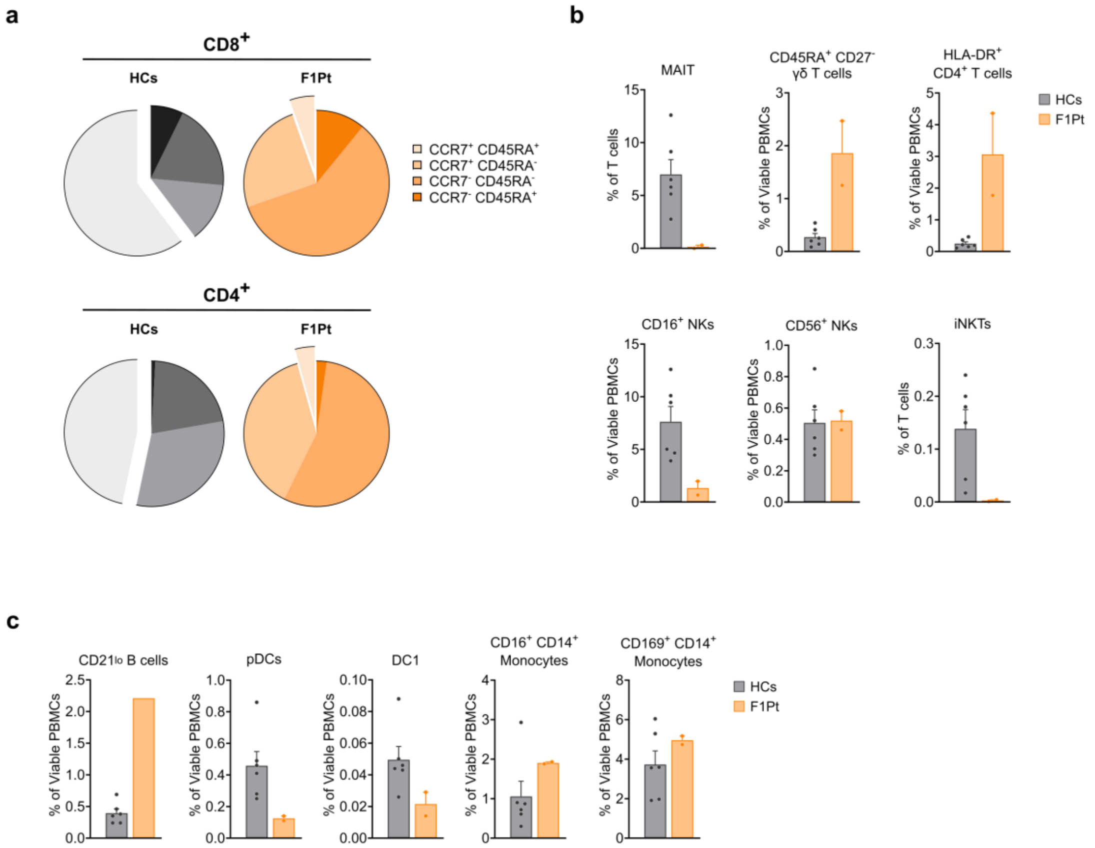


**Supplementary Figure 2.**

**a** Pie charts displaying the distribution of naïve T ($CCR7^+CD45RA^+$), T effector memory ($CCR7^-CD45RA^-$, $T_{EM}$), T central memory ($CCR7^+CD45RA^-$, $T_{CM}$), and terminally differentiated T effector ($CCR7^-CD45RA^+$, $T_{EMRA}$) cells in $CD8^+$ and $CD4^+$ T cells of ATRIP patient (F1Pt) and healthy controls (HCs) (n = 6). **b** Percentages of T and NK subsets in PBMCs of F1Pt and HCs (n = 6), based on manual gating of 25-parameter flow cytometry (FCM) data. Mean and SEM are shown. **c** Percentages of B and innate subsets in PBMCs of F1Pt and HCs (n = 6), based on manual gating of 25-parameter FCM data. Mean and SEM are shown.

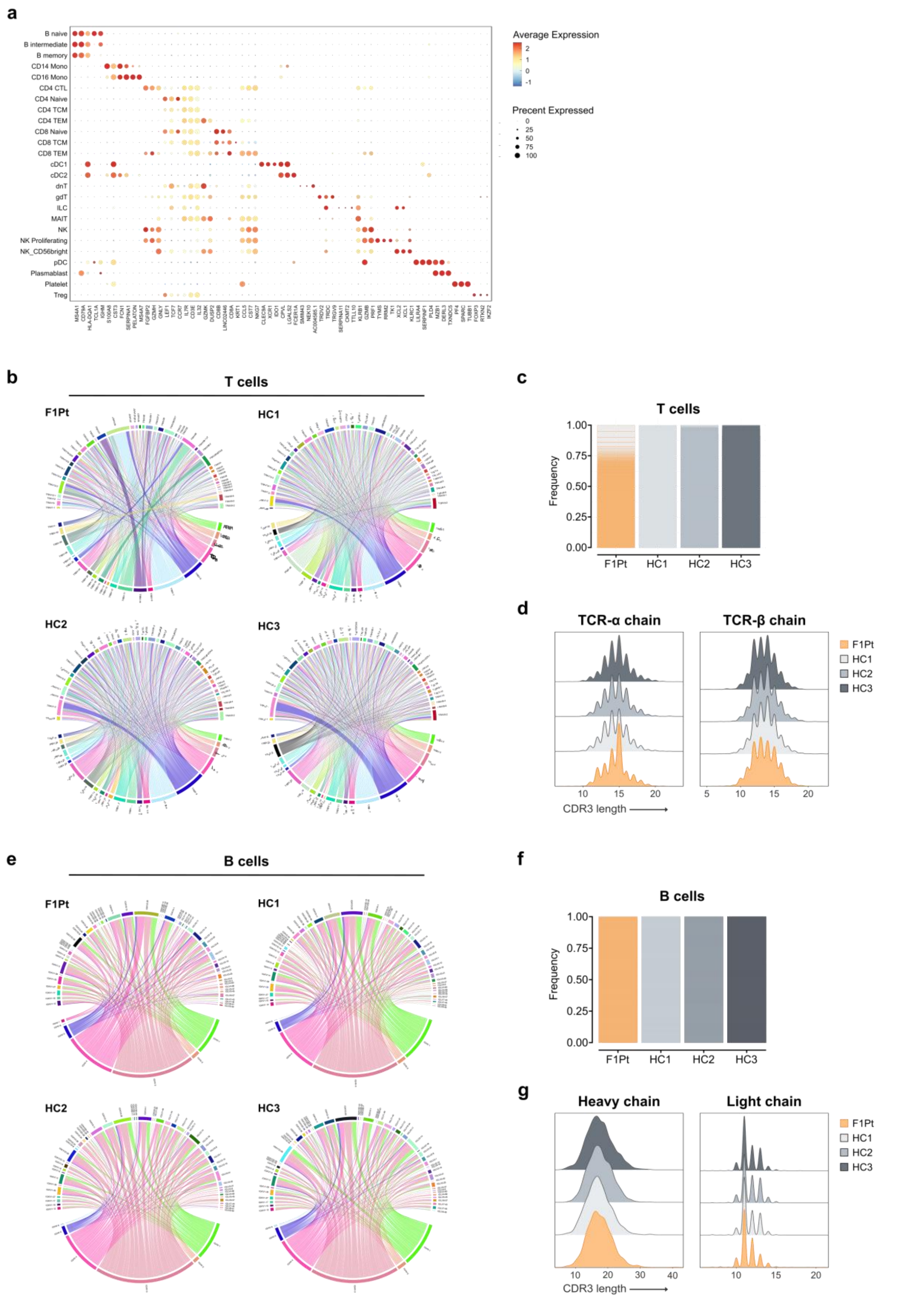


**Supplementary Figure 3.**

**a** Dotplot depicting signature genes defining the UMAP clusters shown in Fig. 3a. **b** Circos plots showing the *TRBV* and *TRAV* pairing pattern of T cells of ATRIP patient (F1Pt) and healthy controls (HCs). **c** Frequency of unique T cell clones in F1Pt and HCs. **d** Distribution of the CDR3 region lengths of TCR-α and TCR-β clones of F1Pt and HCs T cells. **e** Circos plots demonstrating the *IGH*, *IGK*, and *IGK* pairing pattern of F1Pt and HCs B cells. **f** Frequency of unique B cell clones in F1Pt and HCs. **g** Distribution of the CDR3 region lengths of heavy and light chain of F1Pt and HCs B cells.

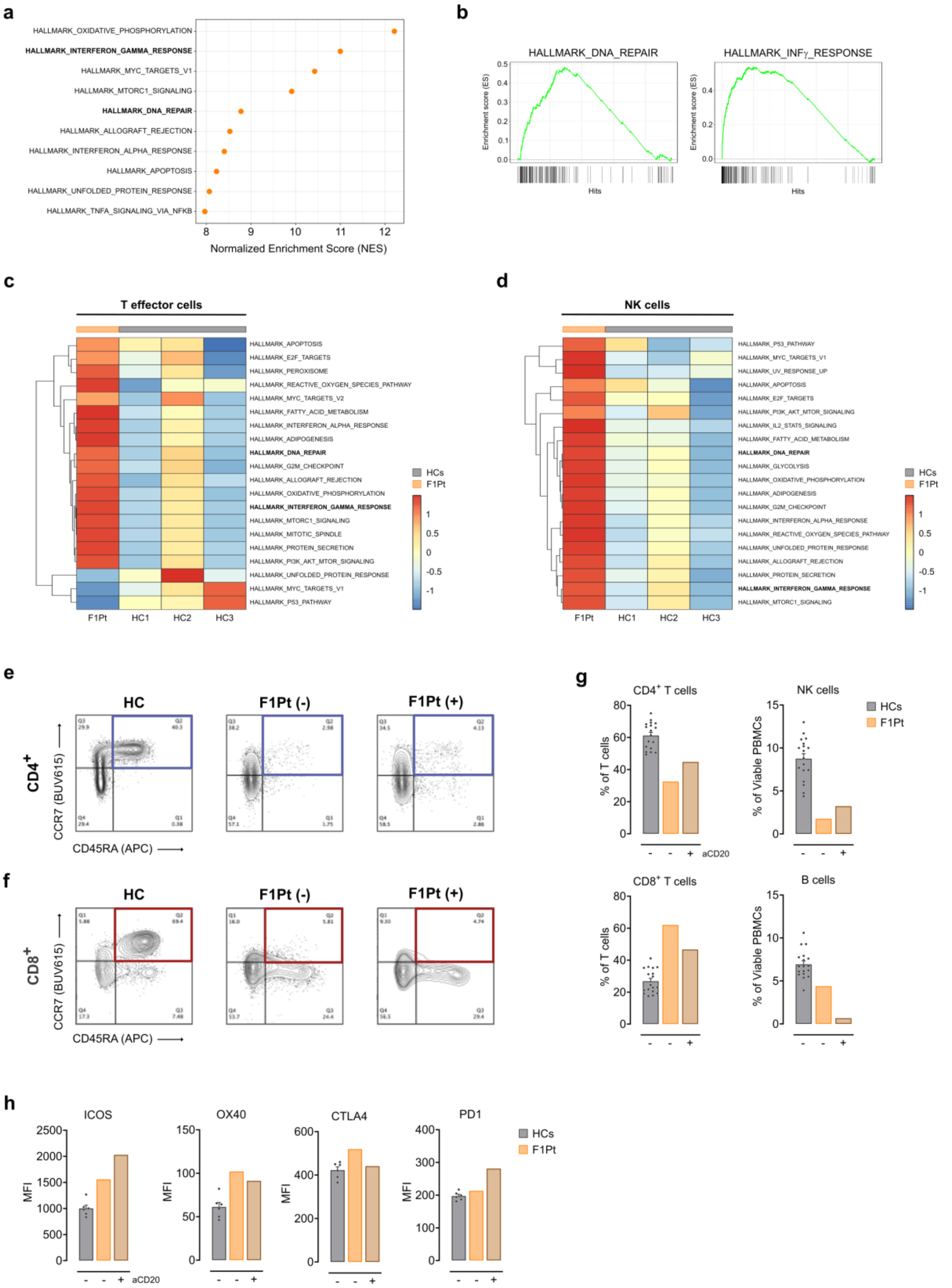


**Supplementary Figure 4.**

**a** MSigDB hallmark gene sets differentially expressed in PBMCs from F1Pt compared to HCs (n = 3). Normalized Enrichment Score (NES) values of the gene sets are depicted. **b** Enrichment plots for two MSigDB hallmark gene sets differentially expressed in PBMCs from F1Pt compared to HCs (n = 3). The profile of the running Enrichment Score (ES) is depicted for both hallmark gene sets. **c** Heatmap displaying the top 20 enriched hallmark gene sets (MSigDB) in T effector cells of F1Pt compared to HCs (n = 3). **d** Heatmap showing the top 20 enriched altered hallmark gene sets (MSigDB) in NK cells of F1Pt compared to HCs (n = 3). **e** Contour plot showing $CD4^+$ T cell maturation in HC and F1Pt. F1Pt (-) represents pre-treatment with anti-CD20 mAb (aCD20), F1Pt (+) represents post-treatment with anti-CD20. **f** Contour plot displaying $CD8^+$ T cell maturation in HC and F1Pt. F1Pt (-) represents pre-treatment with anti-CD20, F1Pt (+) represents post-treatment with anti-CD20. **g** Frequencies of $CD4^+$ T, $CD8^+$ T, NK, and B cells in PBMCs from HCs (n = 18) and F1Pt pre- and post-treatment with anti-CD20. Data represents one experiment, with each datapoint representing one biological replicate. Mean and SEM are shown. **h** ICOS, OX40, PD1, and CTLA4 expression on $CD4^+$ T cells of HCs (n = 6) and F1Pt pre- and post-treatment with anti-CD20 mAb. Bar plots display median fluorescence (MFI). Mean and SEM are shown. Source data are provided as a Source Data file.

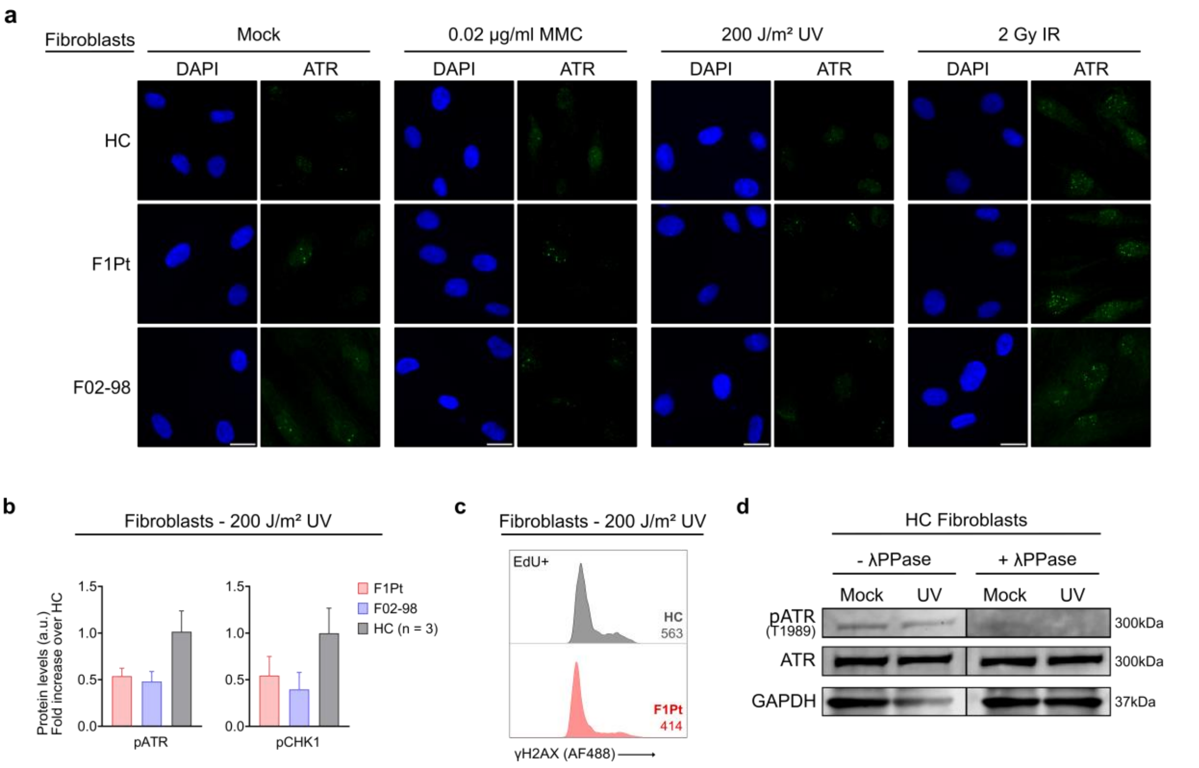


**Supplementary Figure 5.**

**a** Representative immunofluorescence images of cells shown in Fig. 2a with DAPI and ATR staining. Fibroblasts from a healthy control (HC), ATRIP patient (F1Pt), and ATR patient (F02-98) were left untreated or exposed to 0.02 µg/ml Mitomycin C (MMC), 200 J/m² UV or 2 Gy IR. ATR was stained by immunofluorescence following 24h (MMC) or 3h (UV and IR) of treatment. Images are representative of three independent experiments. Scale bars are 20 µm. **b** pATR and pCHK1 levels shown in Fig. 5b were quantified and represent three independent experiments. Bar graph depicts pATR and pCHK1 levels post 200 J/m² UV treatment, expressed as a fold increase over the mean levels observed in three healthy controls. Mean and SD are depicted. **c** γH2AX expression was determined by flow cytometric analysis 3h following 200 J/m² UV exposure in EdU+ fibroblasts of HC and F1Pt. Median fluorescence intensity (MFI) is annotated on the histogram. Data are reflective of one experiment. **d** Immunoblotting of T1989-pATR and total ATR with and without lambda phosphatase (λPPase) treatment on HC fibroblasts, untreated or 3h after 200 J/m² UV exposure. GAPDH serves as a loading control. Source data are provided as a Source Data file.

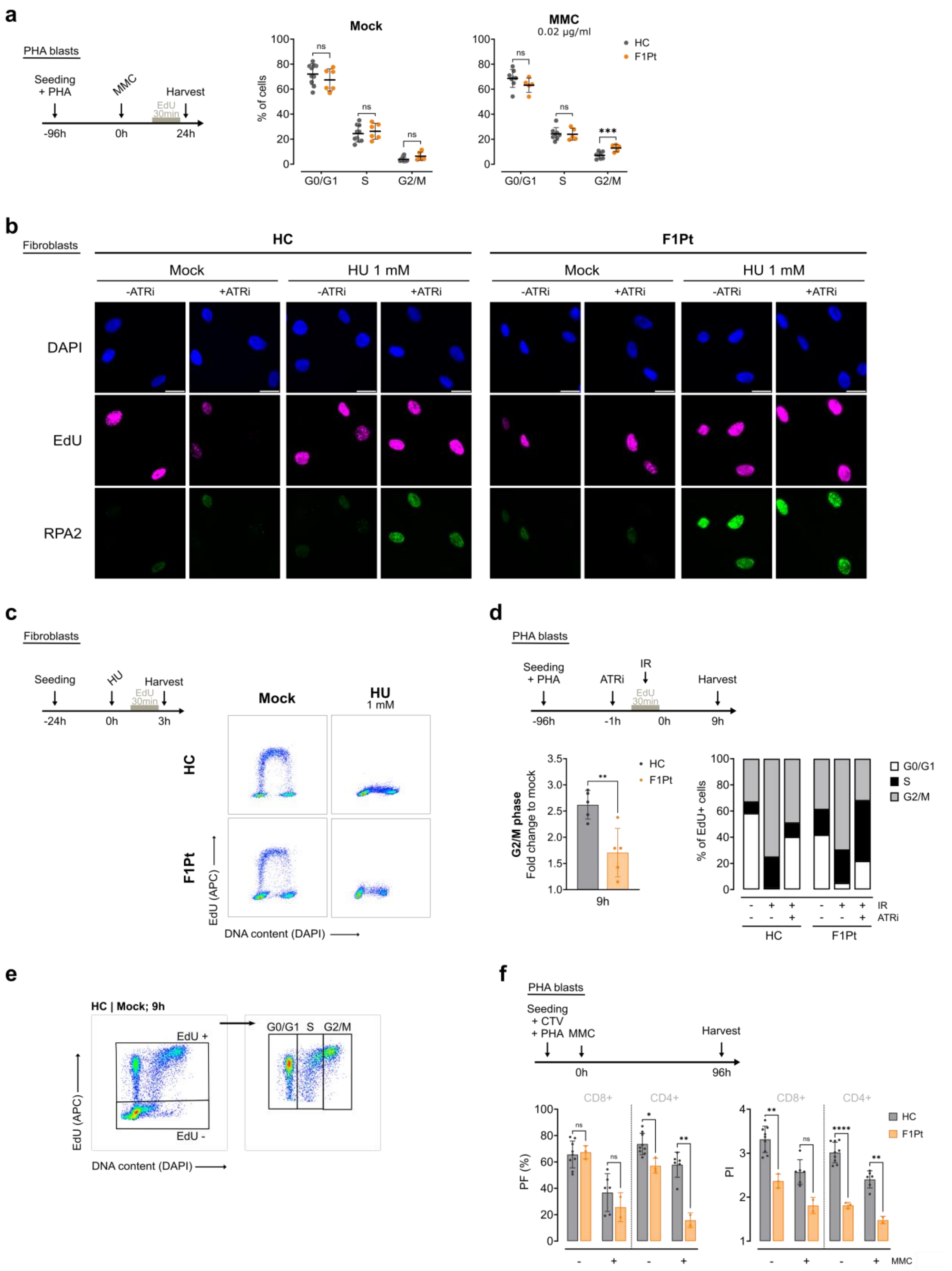


**Supplementary Figure 6.**

**a** Cell cycle distributions of PHA blasts from healthy controls (HCs) and ATRIP patient (F1Pt). Cells were either untreated or treated with 0.02 µg/ml Mitomycin C (MMC) and subsequently harvested at the timepoints indicated on the schematics. Scatter dot plot depicts data from at least five independent experiments. Mean and SD are shown. ns: not significant, *** p<0.001 (multiple paired t-tests). **b** Representative immunofluorescence images with DAPI, EdU, and RPA staining of data shown in Fig. 6d. HC and F1Pt fibroblasts were untreated or exposed to 1 mM hydroxyurea (HU) for 3h. Images are representative of three independent experiments. Scale bars are 20 µm. **c** Flow cytometric (FCM) EdU pulse-labeling profiles of HC and F1Pt fibroblasts demonstrating the inhibiting effect of HU treatment on replication progression. Cells were exposed to 1 mM HU for 3h and subsequently harvested. Data are representative of two experiment. **d** EdU pulse-chase analysis after exposure to 4 Gy IR in the absence or presence of 20 nM ATRi. HC and F1Pt PHA blasts were harvested after 9h. Bar plot (left) shows EdU+ cells present in G2/M phase after IR exposure, depicted as a fold change over the percentage observed in the mock condition. Data represents 5 independent experiment. Mean and SD are shown. **p<0.01 (two-tailed paired t-test). Bar plot (right) shows percentages of EdU+ cells present in G0/G1, S, and G2/M phase. Data from one experiment is shown. **e** FCM gating strategy of EdU pulse-chase kinetics presented in Fig. 6g and Supplementary Fig. 6d. **f** Precursor frequency (PF) and proliferation index (PI) of CD8$^+$ and CD4$^+$ PHA blasts from HC and F1Pt. Cells were labeled with CellTrace Violet (CTV) and subsequently cultured for 96h in the presence or absence of 0.02 µg/ml MMC. Data of at least two independent experiment is shown, with each datapoint representing one biological replicate. Mean and SD are shown. ns: not significant, *p<0.05, **p<0.01, **** p<0.0001 (multiple paired t-tests). Source data are provided as a Source Data file.

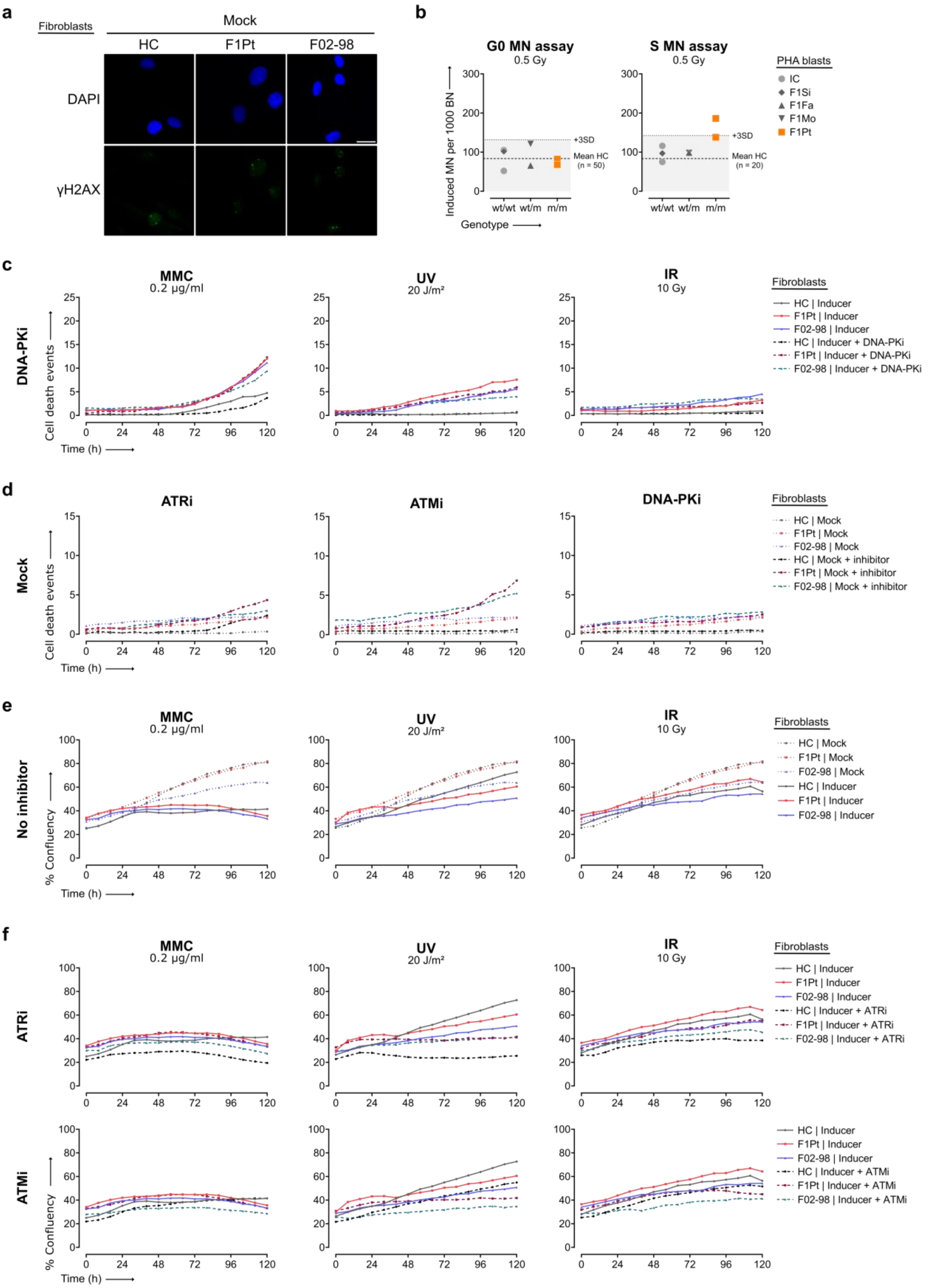


**Supplementary Figure 7.**

**a** Representative immunofluorescence images with DAPI and γH2AX staining of data shown in Fig. 7A. γH2AX foci are shown in untreated fibroblasts from a control (HC), ATRIP patient (F1Pt), and ATR patient (F02-98).

Images are representative of three independent experiments. Scale bars are 20 µm. **b** Using the G0 and S micronucleus (MN) assay, micronuclei were scored in PHA blasts exposed to 0.5 Gy ionizing radiation (IR). Mean MN values of a reference HC group are indicated by dashed lines. Dotted lines correspond to the mean of HCs + 3SD and serve as a cut-off for sensitivity to IR. Data of two independent experiments is shown for F1Pt, in each experiment an internal control (IC) was included. One experiment was performed for the family members (F1Si, F1Fa, F1Mo). **c, d** Fibroblasts from HC, F1Pt, and F02-98 were exposed to genotoxic inducers (0.2 µg/ml MMC, 20 J/m$^2$ UV, or 10 Gy IR) in combination with a specific kinase inhibitor (2 µM DNA-PKi) (**c**) or only treated with 20 nM ATRi, 10 µM ATMi, or 2 µM DNA-PKi (**d**). Cell death was monitored by live imaging up to 120h by quantifying the number of cells stained with SYTOX Green. Represented cell death kinetic plots are reflective of at least two independent experiments. **e, f** Corresponding proliferation curves of the data shown in Fig. 7g and h. Fibroblasts from HC, F1Pt, and F02-98 were untreated or exposed to genotoxic inducers (0.2 µg/ml MMC, 20 J/m$^2$ UV, or 10 Gy IR) without (**e**) or with (**f**) specific kinase inhibitors (20 nM ATRi or 10 µM ATMi) as indicated. Percentage of confluency was monitored by live imaging up to 120h. Data is representative of at least two independent experiments. Source data are provided as a Source Data file.

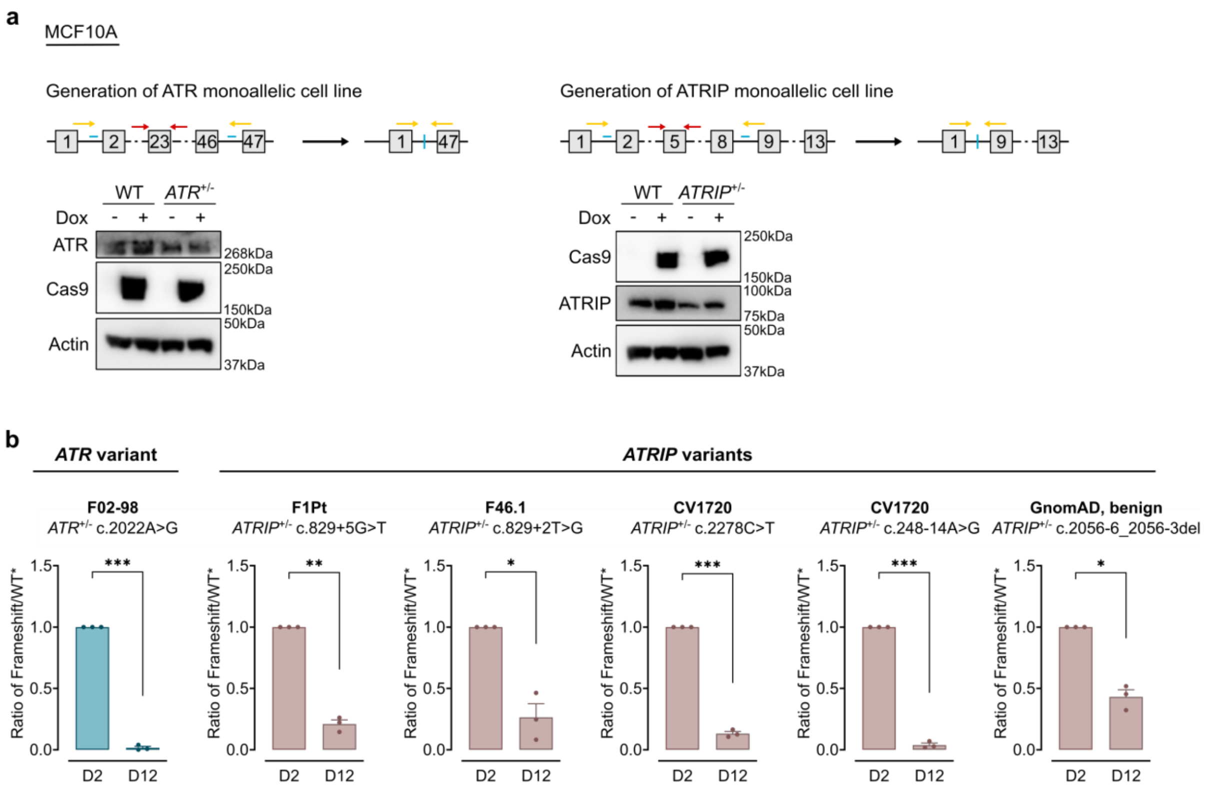


**Supplementary Figure 8.**

**a** Schematics of genome editing of the *ATR* and *ATRIP* allele (top). Blue lines represent the location of crRNAs, orange arrows depict the joint primers annealing to sequence outside the crRNA cutting regions, and red arrows represent the primers annealing to depletion region, used for WT allele detection. The western blot verification of mono-allelic cells is shown (bottom). Cells were seeded in 6 cm dish and 1 µg/ml doxycycline was added in the culture medium 24 hours before cell lysis to induce the expression of cas9 protein. **b** Cell fitness of MCF10A mono-allelic cells with frameshift mutations serving as internal control. Each variant has three independent biological replicates. Mean and SEM are shown. *p<0.05, **p<0.01, ***<0.001 (two-tailed paired t-tests). Source data are provided as a Source Data file.

### Supplementary Tables

**Supplementary Table 1. Clinical features of ATRIP patient (F1Pt).** The clinical presentation is compatible with microcephaly primordial dwarfism (MPD). *Personally identifiable patient information was redacted in accordance with medRxiv requirements.*

**Supplementary Table 2**. **Scores indicating aberrant splicing as predicted by various *in silico* tools (GRCh38, hg38).**

| Prediction tool | g.48457421G>T, c.829+5G>T | g.48457418T>G, c.829+2T>G |
|---|---|---|
| SpliceAI Delta Score (donor loss) | 0.71 | 0.73 |
| Pangolin Delta Score (splice loss) | 0.75 | 0.79 |
| RF Score | 0.874 | 0.926 |
| ADA Score | 0.99909 | 0.99997 |
| MaxEntScan Difference | 3.827 | 7.647 |
| CADD/PHRED Score | 23.5 | 33.0 |

**Supplementary Table 3. Overview of homozygous *ATRIP* splice variants in gnomAD v4.0.0.**

| GnomAD variant ID (GRCh38) | rsID (dbSNP) | HGVS nomenclature (NM_130384.3) | Allele frequency (gnomAD v4.0.0) | Number of homozygotes (gnomAD v4.0.0) | SpliceAI Delta Score (effect, location) | ClinVar (Accession) | Pangolin Delta Score (effect, location) |
|---|---|---|---|---|---|---|---|
| 3-48464565-A-C | rs2242150 | c.1975-17A>C | 0.560807635 | 256364 | *0.01 (AG, -88bp)* | Not reported | 0.01 (gain, 17bp**)** |
| 3-48464823-T-C | rs3135937 | c.2056-8T>C | 0.007480602 | 552 | 0.10 (AG, 40bp) | Benign (VCV001267070.6) | 0.02 (loss, 8bp) |
| 3-48464819-C-A | rs3135936 | c.2056-12C>A | 0.007435878 | 549 | 0.05 (AG, 44bp) | Benign (VCV001276028.5) | 0.03 (loss, 12bp) |
| 3-48450188-T-C | rs11922041 | c.381+18T>C | 0.001880775 | 45 | *0.00 (AL, -151bp)* | Benign (VCV001621536.5) | 0.00 (loss, -18bp) |
| 3-48450019-T-A | rs182766845 | c.248-18T>A | 0.004657135 | 26 | 0.02 (AG, 2bp)<br>0.02 (AL, 18bp) | Benign (VCV001613091.5) | 0.11 (loss, 18bp) |
| 3-48464149-C-T | rs138467436 | c.1974+17C>T | 0.000717299 | 11 | 0.00 (DL, -17bp)<br>*0.04 (DG, 119bp)* | Benign (VCV001990326.1) | NA |
| 3-48464815-A-AC | rs201468664 | c.2056-9dup | 0.001302093 | 2 | 0.05 (AG, 48bp) | Benign (VCV001987555.1) | 0.04 (loss, 16bp) |
| 3-48464678-C-T | NA | c.2055+16C>T | 0.000001368 | 1 | 0.00 (DG, -16bp) | Not reported | 0.00 (gain, -16bp) |
| 3-48464821-CCTCT-C | rs754954019 | c.2056-6_2056-3del | 0.00031091 | 1 | 0.18 (AG, 46bp) | Likely benign (VCV002065669.1) | NA |
| 3-48464822-CT-C | rs367546309 | c.2056-8del | 0.000039899 | 1 | 0.16 (AG, 41bp) | Likely benign (VCV000742744.4) | 0.05 (loss, 1bp) |
| 3-48465474-C-CT | rs765487326 | c.2309-8dup | 0.000013017 | 1 | 0.08 (AG, 15bp) | Not reported | 0.02 (gain, 13bp) |

*AG: acceptor gain; AL: acceptor loss; DG: donor gain; DL: donor loss; NA: not available*

**Supplementary Table 4**. **Considered alternative diagnoses and the associated genes for patient F1Pt.** No other (likely) pathogenic variants were identified in patient F1Pt using whole exome sequencing (WES) analyzing both SNVs and CNVs (MAF cut-off <0.02).

| **Disorder** | **Associated genes** |
|---|---|
| Seckel syndrome | *ATR, RBBP8, CENPJ, CEP152, DNA2, TRAIP, CEP63, NIN, NSMCE2* |
| MOPD type I/III | *RNU4ATAC* |
| MOPD type II | *PCNT* |
| Meier-Gorlin syndrome | *ORC1, ORC4, ORC6, CDT1, CDC6, CDC45L, GMNN, MCM5* |
| Silver-Russell syndrome | *ICR1, IGF2, PLAG1, HMGA2* |
| 3M syndrome | *CUL7, OBSL1, CCDC8* |
| Cornelia de Lange syndrome | *NIPBL, SMC1A, HDAC8, RAD21, SMC3, BRD4* |
| Bloom syndrome | *BLM* |
| Fanconi anemia | *MAD2L2, MAD2B, FANCV, UBE2T, HSPC150, FANCT, PHF9, FANCL, FANCD2, FANCD, FACD, FAD, FANCE, FACE, XRCC2, FANCU, SPGF50, POF17, XRCC9, FANCG, FANCC, FACC, FANCF, BRCA2, FANCD1, BROVCA2, GLM3, PNCA2, RAD51, RECA, MRMV2, FANCR, FANCI, KIAA1794 ,SLX4, BTBD12, MUS312, KIAA1784, KIAA1987, FANCP, ERCC4, XPF, FANCQ, XFEPS, PALB2, FANCN, PNCA3, BROVCA5, RFWD3, FANCW, FANCA, FACA, FA1, FA, FAA, BRCA1, PSCP, BROVCA1, PNCA4, FANCS, RAD51C, FANCO, BROVCA3, BRIP1, BACH1, FANCJ, FANCB, FAAP95, FAAP90, FLJ34064* |

**Supplementary Table 5. Sequences of primers used for variant confirmation (gDNA), splice analysis (cDNA), and real-time qPCR (mRNA) analysis.**

| Primer sequence (5' → 3') | Anneal site | Reference | Comment |
|---|---|---|---|
| ***Variant confirmation (gDNA)*** | | | |
| Forward: *M13*–CTGACAAAGAATGGTAGACATATAATAG<br>Reverse: *M13*–ATTGCTGATCACAGAAATTGG | Exon 5 + intron 5 | ENST00000320211, GRCh38/hg38 (*ATRIP)* | Note, 5' M13 tag to allow subsequent Sanger sequencing with universal M13 primers. Sequence of M13 forward and reverse tag is TGTAAAACGACGGCCAGT and CAGGAAACAGCTATGACC, respectively. |
| ***Splice analysis (cDNA)*** | | | |
| Forward: *M13*–AAGATCACATTTTCTTCTTGAGCA<br>Reverse: *M13*–CTGGGATCAAAGGCTGCTT | Exon 3<br>Exon 7 | ENST00000320211, GRCh38/hg38 (*ATRIP)* | - |
| Reverse: GTGGGGAAGGGACATGTTAG | Exon 5 | ENST00000320211, GRCh38/hg38 (*ATRIP)* | Sanger sequencing primer |
| Forward: GGAAAAACCCTTCTGTGGTT | Exon 5 | | Sanger sequencing primer |
| ***Real-time qPCR analysis (mRNA)*** | | | |
| Hs.PT.58.46500972 | Exon 3-4 | NM_130384 (*ATRIP)* | Primer pair |
| Hs.PT.58.15253191 | Exon 5-6 | | Primer pair |
| ***Housekeeping gene RT-qPCR (mRNA)*** | | | |
| Forward: CTGGAACGGTGAAGGTGACA<br>Reverse: AAGGGACTTCTTGTAACAATGCA | *ACTB*<br>*ACTB* | - | - |

**Supplementary Table 6. Detailed overview of the monoclonal antibodies used for immunophenotyping.**

| Antigen | Fluorochrome | Clone | Company |
|---|---|---|---|
| CD3 | BUV395 | UCHT1 | BD |
| CD8 | BUV496 | RPA-T8 | BD |
| CD278 | BUV563 | DX29 | BD Optibuild |
| CCR7 | BUV615 | 2-L1-A | BD |
| HLA-DR | BUV661 | G46-6 | BD |
| CD95 | BUV737 | 563 | BD Optibuild |
| CD4 | BUV805 | SK3 | BD |
| CXCR3 | BV421 | G025H7 | Biolegend |
| CXCR5 | BV480 | RF8B2 | BD |
| CD16 | BV570 | 3G8 | Biolegend |
| CD152 | BV605 | BNI3 | Biolegend |
| CD39 | BV650 | TU66 | BD Horizon |
| CD25 | BV711 | BC96 | Biolegend |
| CCR4 | BV750 | 1G1 | BD Optibuild |
| CD28 | BV785 | CD28.2 | Biolegend |
| TCRg/d | FITC | B1 | Biolegend |
| PD1 | BB660-P | EH12.1 | BD |
| CD56 | BB790-P | NCAM16-2 | BD |
| CD134 | PerCP Cy5-5 | ACT35 | Biolegend |
| FoxP3 | PE | 206D | Biolegend |
| CCR6 | PE-Dazzle594 | G034E3 | Biolegend |
| SA | PE-Cy5 | 206D | BD |
| CD27 | PE-Cy7 | O323 | Biolegend |
| CD45RA | APC | HI100 | Biolegend |
| Va7.2 | AF700 | 3C10 | Biolegend |
| CD161 | APC-Fire750 | HP-3G10 | Biolegend |
| Va24Ja18 | biotin | 6B11 | Thermofisher |
| CD24 | BUV395 | ML5 | BD |
| CXCR3 | BUV496 | 1C8 | BD Optibuild |
| CCR7 | BUV615 | 2-L1-A | BD |
| HLA-DR | BUV661 | G46-6 | BD |
| CD86 | BUV737 | 2331 (FUN-1) | BD |
| CD3 | BUV805 | UCHT1 | BD |
| CD1c | BV421 | L161 | Biolegend |
| CD185 | BV480 | RF8B2 | BD |
| CD16 | BV570 | 3G8 | Biolegend |
| IgD | BV605 | IA6-2 | Biolegend |
| CD11c | BV650 | 3.9 | Biolegend |
| CD25 | BV711 | BC96 | Biolegend |
| CD14 | BV750 | 63D3 | Biolegend |
| CD20 | BV785 | 2H7 | Biolegend |
| PD1 | BB660 | EH12.1 | BD Horizon |
| CD169 | BB515 | 7-239 | BD Horizon |
| CD123 | BB630 | 7G3 | BD Horizon |
| CD94 | PerCP-Cy5-5 | 18D3 | Biolegend |
| CD21 | PE | Bu32 | Biolegend |
| CD64 | PE-Dazzle594 | 10.1 | Biolegend |
| IgM | PE-Cy5 | G20-127 | BD |
| CD19 | PE-Cy7 | HIB19 | Biolegend |
| CD141 | AF647 | M80 | Biolegend |
| CD27 | R718 | M-T271 | BD |
| CD38 | APC-eFire750 | HIT2 | Biolegend |

**Supplementary Table 7. Designed CRISPR RNAs (crRNAs) and PCR primers for generating the mono-allelic cell lines used in CRISPR-SELECT$^{TIME}$.**

| Mono-allelic cell line | crRNAs | PCR Primers |
|---|---|---|
| ATR$^{+/-}$ | crRNA1:<br>CACCTCTTCAATACACAGGC<br>crRNA2:<br>ATCTTTAATAAGTCTTGTAC | Primer for detecting WT allele:<br>Forward: TGAACCTCACTGTCTTTTCCTT<br>Reverse: AGTTTTAGAGCTGTGAACATCCT<br>Joint Primer:<br>Forward: TGAAGATGTGACCTCCCTCG<br>Reverse: AGGGTTAAGGATGACATGAAGGA |
| ATRIP$^{+/-}$ | crRNA1:<br>TTACTCTTGAGAGCAAAGCA<br>crRNA2:<br>TACAGCCCAATAATGAAACC | Primer for detecting WT allele:<br>Forward: TTGAGCTCAGCAGTTCCAGA<br>Reverse: TCCAGTCCCTGCAGAGATTG<br>Joint Primer:<br>Forward: GAGGGTGAGGCACAAGA<br>Reverse: GGGGAGCACGATGGACTGG |

**Supplementary Table 8. Designed CRISPR RNAs (crRNAs) and PCR primers for variants analysis with CRISPR-SELECT$^{TIME}$.**

| Variants | crRNAs | PCR Primers |
|---|---|---|
| ATR<br>c.2022A>G | ATCCGGGCTAGTTGTGTTAG | Forward:<br>ACACTCTTTCCCTACACGACGCTCTTCCGATCT<br>TCCTTGAGTGGAGAACAGCAGT<br>Reverse:<br>TGACTGGAGTTCAGACGTGTGCTCTTCCGATCT<br>ACCCTGCATACATAGCCAGACA |
| ATRIP<br>c.829+5G>T | ATACATCAATAGATGACTAA | Forward:<br>ACACTCTTTCCCTACACGACGCTCTTCCGATCT<br>ACATGTCCCTTCCCCACCCCTG<br>Reverse:<br>TGACTGGAGTTCAGACGTGTGCTCTTCCGATCT<br>TGTCCCCTCTGTGCCTTGTCAGA |
| ATRIP<br>c.829+2T>G | ATACATCAATAGATGACTAA | Forward:<br>ACACTCTTTCCCTACACGACGCTCTTCCGATCT<br>ACATGTCCCTTCCCCACCCCTG<br>Reverse:<br>TGACTGGAGTTCAGACGTGTGCTCTTCCGATCT<br>TGTCCCCTCTGTGCCTTGTCAGA |
| ATRIP<br>c.2278C>T | GGGGTCAGCATGCTCATCCG | Forward:<br>ACACTCTTTCCCTACACGACGCTCTTCCGATCT<br>GCACGGCCTATCGCAGAAGGAC<br>Reverse:<br>TGACTGGAGTTCAGACGTGTGCTCTTCCGATCT<br>ACCTCTGGAAGAAGCCATGGGGG |
| ATRIP<br>c.248-14A>G | ATTTTTTACAGGTGATCATA | Forward:<br>ACACTCTTTCCCTACACGACGCTCTTCCGATCT<br>CTGCACTCCAACCTGGGTAACA<br>Reverse:<br>TGACTGGAGTTCAGACGTGTGCTCTTCCGATCT<br>TGTGCCTGAAGTACCTCTAATTCGA |
| ATRIP<br>c.2056-6_2056-3del | CGCTCTGACCACCTGAGAGA | Forward:<br>ACACTCTTTCCCTACACGACGCTCTTCCGATCT<br>AGGTGAGTGGGTAGGGGCCAAC<br>Reverse:<br>TGACTGGAGTTCAGACGTGTGCTCTTCCGATCT<br>TCCGCACTGTCAGCCACTGTCT |

**Supplementary Table 9. Designed mutation (MUT) and wild-type (WT*) DNA repair templates for variants analysis with CRISPR-SELECT$^{\text{TIME}}$.**

| Variants | MUT repair template | WT* repair template |
|---|---|---|
| ATR c.2022A>G | CCTGCAGAGCTCCCATGAAGTAATCCGGGC TAGTTGTGTGAGTGGGTTTTTTATCTTATTGC AGCAGCAGAATTCTTGTAACAGAGTTCC | CCTGCAGAGCTCCCATGAAGTAATCCGGGC TAGTTGTGTGAGCGGATTTTTTATCTTATTGC AGCAGCAGAATTCTTGTAACAGAGTTCC |
| ATRIP c.829+5G>T | CAGACGGAGTCAGGATACAAGCCTCTGGTG GGCAGAGAGGGTAATTCCATTACTCATCTAT TGATGTATAACAAGCTACCCAAATATGTG | CAGACGGAGTCAGGATACAAGCCTCTGGTG GGCAGAGAAGGTAAGTCCATTACTCATCTAT TGATGTATAACAAGCTACCCAAATATGTG |
| ATRIP c.829+2T>G | TGCCAGACGGAGTCAGGATACAAGCCTCTG GTGGGCAGAGAGGGGAAGTCCATTACTCAT CTATTGATGTATAACAAGCTACCCAAATAT | TGCCAGACGGAGTCAGGATACAAGCCTCTG GTGGGCAGAGAAGGTAAGTCCATTACTCATC TATTGATGTATAACAAGCTACCCAAATAT |
| ATRIP c.2278C>T | TGCATCAGTTTGACCAGGTGATGCCGGGGG TCAGCATGCTCATTTGAGGGCTTCCTGATGT GACGGACTGTGAAGGTAAGCCTGCCAGAG | TGCATCAGTTTGACCAGGTGATGCCGGGGG TCAGCATGCTCATTCGTGGGCTTCCTGATGT GACGGACTGTGAAGGTAAGCCTGCCAGAG |
| ATRIP c.248-14A>G | TCTGGCAAAAGTGGGAAAATATTGGGTCCTG AAATGTATATGGAGCTATTTTTTACAGGTGAT CACAAGGTCCACAGATTATTAGATGGC | TCTGGCAAAAGTGGGAAAATATTGGGTCCTG AAATGTATATGGAACTATTTTTTACAGGTGAC CACAAGGTCCACAGATTATTAGATGGC |
| ATRIP c.2056-6_2056-3del | TAGGGTGCAGGCCATGGTGGCACCAGGCCT CAGTCTGCACCCCCCCTCAGGTGGTCAGAG CGCTCACGGTGATGTTGCACAGACAGTGGC | GTGCAGGCCATGGTGGCACCAGGCCTCAGT CTGCACCCCCCCTCTCTCAGGTCGTCAGAG CGCTCACGGTGATGTTGCACAGACAGTGGC |
